# Understanding the impact of mobility on COVID-19 spread: a hybrid gravity-metapopulation model of COVID-19

**DOI:** 10.1101/2022.12.16.22283600

**Authors:** Sarafa A. Iyaniwura, Notice Ringa, Prince A. Adu, Sunny Mak, Naveed Z. Janjua, Michael A. Irvine, Michael Otterstatter

## Abstract

The outbreak of the severe acute respiratory syndrome coronavirus 2 started in Wuhan, China, towards the end of 2019 and spread worldwide. The rapid spread of the disease can be attributed to many factors including its high infectiousness and the high rate of human mobility around the world. Although travel/movement restrictions and other non-pharmaceutical interventions aimed at controlling the disease spread were put in place during the early stages of the pandemic, these interventions did not stop COVID-19 spread. To better understand the impact of human mobility on the spread of COVID-19 between regions, we propose a hybrid gravity-metapopulation model of COVID-19. Our model explicitly incorporates time-dependent human mobility into the disease transmission rate, and has the potential to incorporate other factors that affect disease transmission such as facemasks, physical distancing, contact rates, etc. An important feature of this modeling framework is its ability to independently assess the contribution of each factor to disease transmission. Using a Bayesian hierarchical modeling framework, we calibrate our model to the weekly reported cases of COVID-19 in thirteen local health areas in metro Vancouver, British Columbia (BC), Canada, from July 2020 to January 2021. We consider two main scenarios in our model calibration: using a fixed distance matrix and time-dependent weekly mobility matrices. We found that the distance matrix provides a better fit to the data, whilst the mobility matrices have the ability to explain the variance in transmission between regions. This result shows that the mobility data provides more information in terms of disease transmission than the distances between the regions.

## 1 Introduction

The pandemic of the severe acute respiratory syndrome coronavirus 2 (SARS-CoV-2), which started in the city of Wuhan, Hubei province, China [64] has since spread all over the world with over 585 million reported cases and 6.4 million reported deaths, as of August 2022 [63]. In human populations, the virus can be transmitted through the inhalation of infectious droplets in aerosols, exposure to infectious respiratory fluids, coughing, sneezing, and having physical contact with an infected individual. It can also be transmitted indirectly when a susceptible individual comes in contact with a contaminated surface, such as door handles or other commonly shared surfaces or objects [37, 47, 21, 65]. SARS-CoV-2 is the casual agent for the coronavirus disease 2019 (COVID-19), and is estimated to be more infectious compared to other coronaviruses such as the severe acute respiratory syndrome (SARS) and the Middle East respiratory syndrome coronavirus (MERS) [44, 1]. The COVID-19 disease was declared a public health emergency by the World Health Organization (WHO) on January 20, 2020 [66] and a pandemic on March 11, 2020 [67].

Due to the fast spread of COVID-19, during the early stages of the pandemic, governments around the implemented non-pharmaceutical interventions (NPIs) such as movement/travel restrictions, wearing of facemasks, closure of schools and business, physical distancing, etc. [48, 40, 20, 19, 7], to limit the spread of the disease. Although, the implementation of these NPIs helped in slowing down the spread of COVID-19, the disease still continues to spread under these restrictions. In addition, these NPIs have significant social and economic effects around the world [9, 14, 32], and could not be put in place for too long. The development of safe and effective COVID-19 vaccines brought some relief and were introduced to replace stringent NPIs [41, 51]. The first set of COVID-19 vaccines became available towards the end of 2020 [18]. These vaccines provide significant protection against the earlier strains of SARS-CoV-2 virus [58, 49, 50]. However, the emergence of highly infectious mutant strains such as Omicron variant led to the continuous spread of the disease.

The first case of COVID-19 was reported in Wuhan, China, in December 2019 [29]. On the 12^*th*^ of January 2020, the first case of the disease outside of China was confirmed in Thailand [31]. By January 30, 2020, COVID-19 has spread to 18 countries outside of China with a total of 7,818 confirmed cases worldwide [31]. The first confirmed case of COVID-19 in Africa was reported on February 14, 2020 [30, 26], in North America, January 21, 2020 [2], and in Europe, January 24, 2020 [53]. COVID-19 has spread more rapidly and widely around the world than previous outbreaks of coronaviruses. This spread can be attributed to globalization, settlement and population characteristics, and high human mobility [52]. Several studies have looked at the effect of human mobility on the spread of COVID-19 [38, 27, 22]. In Kraemer, Moritz UG, et al [38], real-time human mobility data was used to investigate the role of case importation in the spread of COVID-19 across cities in China. The impact of human mobility network on the onset of COVID-19 in 203 countries was studied in [27]. They used exponential random graph models to analyze country-to-country spread of the disease. Their study suggested that migration and tourism inflow contributed to COVID-19 case importation, and that a mixture of human mobility and geographical factors contribute to the global transmission of COVID-19 from one country to another. Human mobility data collected via mobile devices such as cell phones, smartwatches, e-readers, tablets etc., has also been used to study the spread of COVID-19 [15, 36, 46, 68]. In [15], county-level cell phone mobility data collected over a period of 1 year in the US was used to study the spatio-temporal variation in the relationship between COVID-19 infection and mobility. They found that in the spring 2020, sharp drop in mobility often coincide with decrease in COVID-19 cases in many of the populous counties.

Mathematical models have been used to study the relationship between the spatio-temporal spread of COVID-19 and human mobility [68, 70, 3, 8, 72, 5, 12, 25, 62]. A city-based epidemic and mobility model together with multi-agent network technology and big data on population migration were used to simulate the spatio-temporal spread of COVID-19 in China [62]. In [10], a stochastic, data-driven metapopulation model was used to study the initial wave of COVID-19 in Belgium, and also to study different re-opening strategies. Their model incorporates the mixing and mobility of different age groups in Belgium. Another stochastic metapopulation model was used to study the spread of COVID-19 in Brazil [12]. This model assumes that epidemics start in highly populated central regions and propagate to the countrysides. For many states, they found strong correlations between the delay in epidemic outbreaks in the countrysides and their distance from central cities. In [13], an SEIR country-wide metapopulation model was used to study the spread of COVID-19 in England and Wales. The model was used to predict the COVID-19 epidemic peak in England and Wales, and also to study the effect of different non-pharmaceutical intervention strategies on the predicted epidemic peaks. Similarly, in [45] a stochastic SIR model was applied to describe the spatiotemporal spread of COVID-19 across 33 provincial regions in China and to also evaluate the effectiveness of various local and national intervention strategies. Their model incorporates an outflow mobility index for all the regions and the proportion of travelers between regions. More discussions on human mobility and COVID-19 transmission can be found in the systematic review article [71]. The relative contribution of mobility data to the observed variance in the COVID-19 transmission rates between regions still remains an unexplored problem.

Here, we develop a hybrid gravity-metapopulation modeling framework for studying the spread of COVID-19 within and between different regions. Our modeling framework allows for explicit incorporation of factors that affect disease transmission such as human mobility, facemasks, physical distancing, contact rate, etc., into a time-dependent disease transmission rate. Spatial mobility data collected through mobile device counts are used to infer influx into a region and also to quantify movement within the region. As an illustration, we use Bayesian hierarchical modeling framework to calibrate our model to the weekly reported cases of COVID-19 in the thirteen local health areas (LHAs) of Fraser health authority (Fraser Health), British Columbia (BC), Canada, from July 2020 to January 2021. The study area comprises of 1.9 million population on the eastern sections of the Greater Vancouver area. We estimate region-specific scaling parameters for computing baseline disease transmission rates for each region, and a parameter for quantifying the contribution of mobility to disease transmission. In addition, we estimate a time-dependent piece-wise constant scaling parameter to account for the cumulative effect of the remaining factor that affect disease dynamics, which are not explicitly included in our model. We consider two main model structures in our example, which are determined by the mobility matrices used: one with a distance matrix (computed using the distances between the regions, based on the population weighted centroid) and another with time-dependent mobility matrices computed from mobile device data. The results from these two scenarios are used to test the hypothesis of whether the time-dependent mobility matrices, computed from mobile device counts, provide more information about human mobility, with respect to disease transmission between the regions than the distances between the region.

## 2 Methods

### 2.1 Mathematical Model

We develop a hybrid gravity-metapopulation model to study the dynamics of COVID-19, within and between regions. The model stratifies the population of each region into six compartments: susceptible (S), exposed (E), pre-symptomatic infectious (P), symptomatic infectious (*I*_1_ and *I*_2_), and recovered (R). Individuals in the pre-symptomatic infectious compartment are infectious (can transmit the disease) but do not show symptoms yet. Similar to [35, 11], we divided the infectious compartment into two classes so that the recovery time follows a Gamma distribution rather than an exponential distribution. This way, a symptomatic infectious individual spends the first half of their infectious period in *I*_1_ and the other half in *I*_2_. Our model assume that there are no reinfections.

A schematic diagram of the model illustrated for four (4) regions is shown in Figure 1, where the gray circles on the left represent the regions, while the black arrows show the interactions and movements of individuals between the regions. On the right, we have an illustration of the population dynamics in each of the regions, where the subscript *j* represents the *j*^th^ region. The black arrows here show the transition of individuals through the different stages of COVID-19 at the rates indicated beside the arrows. The red dashed arrows indicate disease transmission. Observe that there is a red dashed arrow extending from each of the remaining three regions into region *j*, these arrows account for the contributions of infectious individuals in the three regions to disease transmission in the *j*^th^ region. The ordinary differential equations (ODEs) for the model are given by (see Figure 1 for model schematic diagram and definition of state variables):

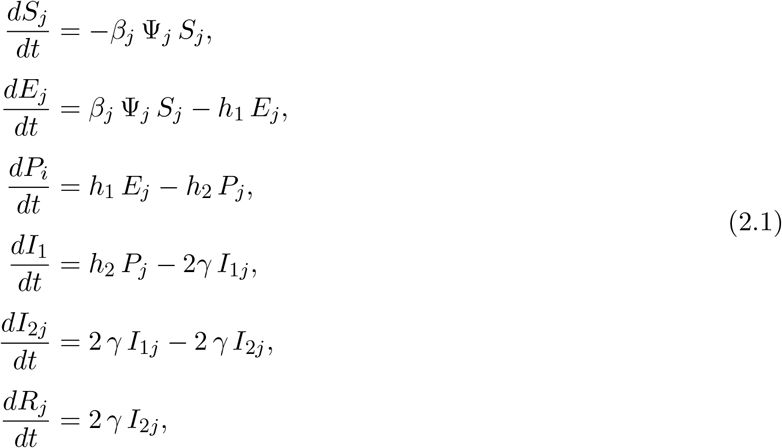

where *β*_*j*_ ≡ *β*_*j*_(*t*) is the time-dependent disease transmission rate for region *j* defined by

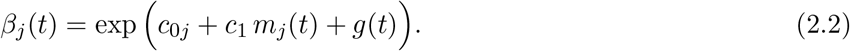

Here, *c*_0*j*_ is the scaling parameter for the baseline disease transmission rate for region *j, c*_1_ is the scaling parameter used to remove biases from the time-series mobility data, and *g*(*t*) is a time-dependent piece-wise parameter used to account for other factors that affect disease transmission other than human mobility (e.g. facemask, social distancing, contact rates, etc.), which are not explicitly incorporated into the model. Movement within the *j*^th^ region is captured by a time-series mobility data represented by *m*_*j*_(*t*). This data is used as a proxy for the time-dependent contact rate in the region. We observe from (2.2) that 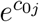 is the baseline disease transmission rate for region *j*, while 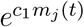 incorporates the effect of human mobility into the transmission rate. Lastly, *e*^*g*(*t*)^ is the effect of other factors that affect disease transmission, which are not explicitly incorporated into the model, on the disease spread. Although, the formulation in (2.2) explicitly incorporates only human mobility into the disease transmission rate, this formulation can be extended to include other factors that affect disease transmission such as facemaks, physical distancing, etc. See more details in the discussion section.

**Figure 1:**
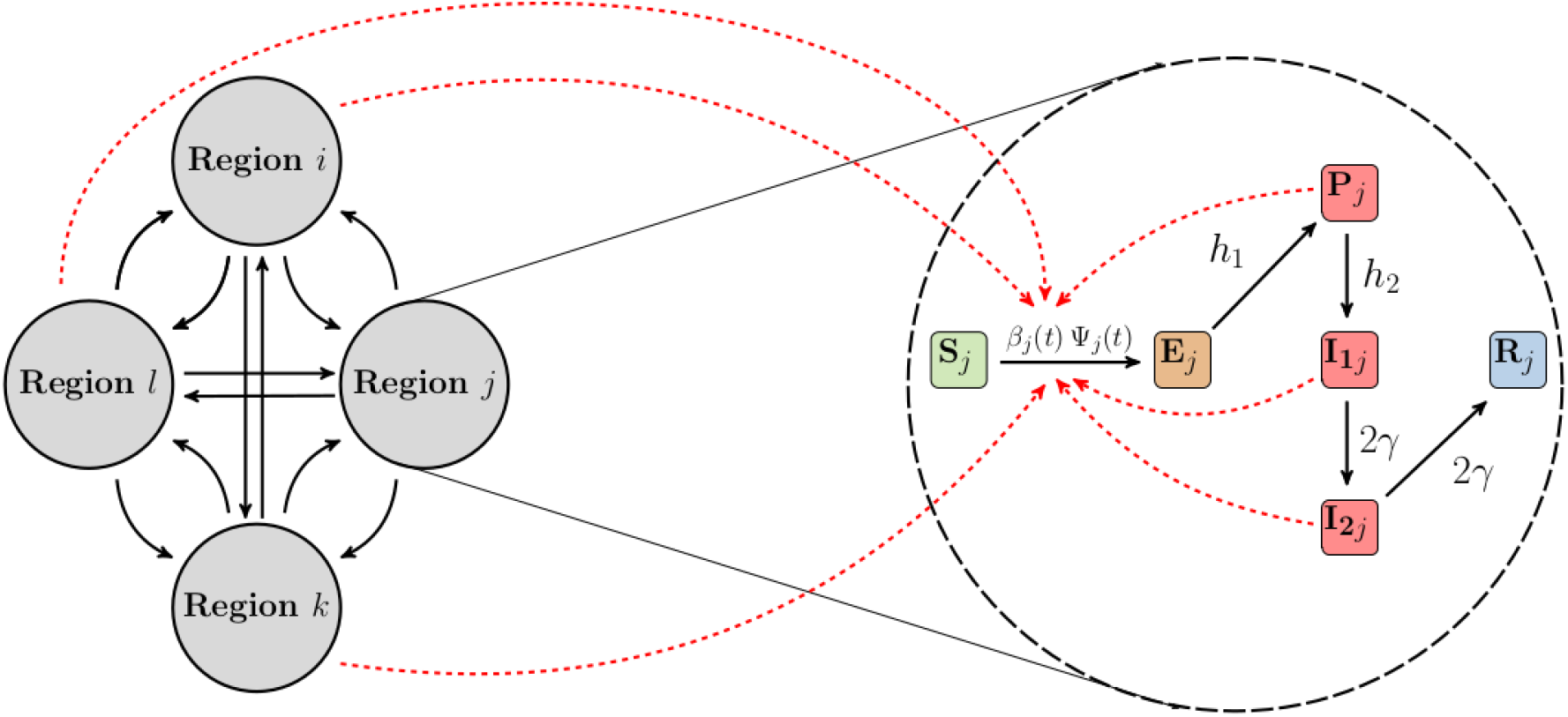
Schematic of the model. An illustration of our hybrid gravity-metapopulation model for four regions. Interactions and movements between the regions (left) and stratification of the population of each region based on disease stages (right). Model compartments are defined as follows: Susceptible (S_j_); exposed (E_j_); pre-symptomatic infectious (P_j_); symptomatic infectious (I_1j_ and I_2j_); and recovered (R_j_) for region j. Our model assumes that there are no reinfections. The black arrows show the movement of individuals from one region to another (left) and the transition of individuals through the different stages of COVID-19 at the rates indicated beside the arrows (right). The red dashed arrows indicate disease transmission (see (2.1) for more details).

The parameter Ψ_*j*_ ≡ Ψ_*j*_(*t*) in (2.1) is used to incorporate infectious interactions within the *j*^th^ region, and their contribution to disease transmission in the region. In terms of a homogeneous single population model, this parameter would represent the probability of making an infectious contact in the population. Here, Ψ_*j*_ is defined as

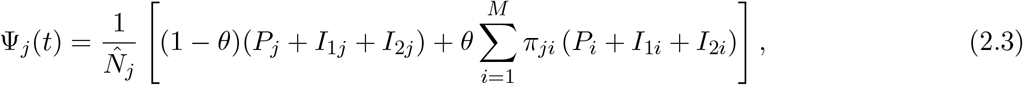

where 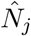 is the adjusted total population in region *j*, which incorporates the increase in the population of the region due to the influx from other regions, and it is given by

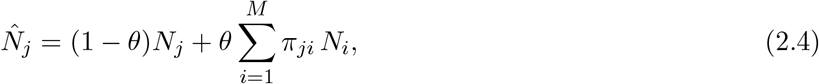

where *M* is the total number of regions under consideration and *N*_*j*_ is the baseline population size of the *j*^th^ region. The parameter *θ* measures the effective contribution of human mobility to disease transmission in all the regions. In (2.3) and (2.4), *π*_*ji*_ is the probability that an individual who migrated into region *j*, originated from region *i*, given that he/she is from one of the other regions under consideration. We compute this probability using two different approaches. The first approach uses the distances between the regions. In this case, *π*_*ji*_ is given by

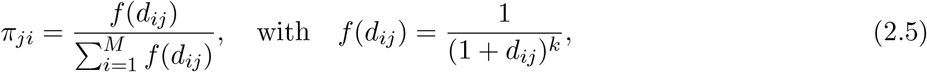

Where *d*_*ij*_ ≡ *d*_*ji*_ is the distance from region *i* to region *j, k* ∈ ℝ^+^ and *M* is the total number of regions considered. The second approach used to compute the probability *π*_*ji*_ involves using mobile device data (see §2.2 for details).

The model parameters, their descriptions, and values are provided in Table 1. The estimated parameters are presented in the result section (§3). As an illustration of concept, we consider the thirteen (13) local health areas (LHA) of Fraser health authority, British Columbia (BC), Canada. These regions include the communities of Abbotsford, Agassiz/Harrison, Burnaby, Chilliwack, Delta, Hope, Langley, Mission, Maple Ridge/Pitt Meadows, New Westminster, South Surrey/White Rock, Surrey and Tri-Cities. Fraser health authority (Fraser Health) is the largest of the five regional health areas in BC, with 12 acute care hospitals and providing health care to over 1.9 million people [28]. It has a width of 150 km. Figure 2 shows a map of British Columbia (left) and Fraser Health (right) with the 13 health regions shaded in different colors. The population sizes of the regions are detailed in Table S3 of Supplementary material A.5. We use a Bayesian hierarchical modeling framework to calibrate our model to the weekly reported cases of COVID-19 in these 13 LHAs, from July 2020 to January 2021. From the model calibration, we estimate the parameters *c*_0*j*_, *c*_1_ and *g*(*t*), which are used to construct and study the time-dependent disease transmission rate for each region, and to study the dynamics of the time-dependent piece-wise parameter *g*(*t*). We also estimated the parameter *θ*, used to quantify the effect of mobility, both within and between the regions, on disease transmission in the regions

**Table 1:** Model parameters, their description, and value. The estimated parameters are presented in the result section (§3). The population sizes for the regions are shown in Figure 2 and Table S3 of Supplementary material A.5.

| Parameter | Description | Value | References |
| --- | --- | --- | --- |
| $\beta_j(t)$ | Time-dependent disease transmission rate for region $j$ | — | Computed using (2.2) |
| $c_{0j}$ | $\exp(c_{0j})$ is the baseline disease transmission rate for region $j$ | — | Estimated |
| $c_1$ | Scaling parameter for the time-series mobility data | — | Estimated |
| $g(t)$ | Time-dependent piece-wise scaling parameters used to account for the effect of other factors that affect disease transmission | — | Estimated |
| $h_1$ | Rate of transitioning from exposed to pre-symptomatic infectious | 1/5 (days<sup>-1</sup>) | [39, 4, 73] |
| $h_2$ | Rate of transitioning from pre-symptomatic infectious to symptomatic infectious | 1 (days<sup>-1</sup>) | [42, 59, 23, 4] |
| $\gamma$ | Infection recovery rate | 1/5 (days<sup>-1</sup>) | [59, 23, 4] |
| $\theta$ | Measures the effect of mobility on disease transmission | — | Estimated |
| $d_{ij}$ | Distance from region $i$ to region $j$ | See Figure 3 | Computed using [55] |
| $\pi_{ji}$ | Probability that an individual moving in region $j$ is from region $i$ | See Figure 3 & 4 | Computed using (2.5) |
| $N_j$ | Baseline population size for region $j$ | See Table S3 | [6] |
| $\hat{N}_j$ | Adjusted population size for region $j$ | — | Computed using (2.4) |

**Figure 2:**
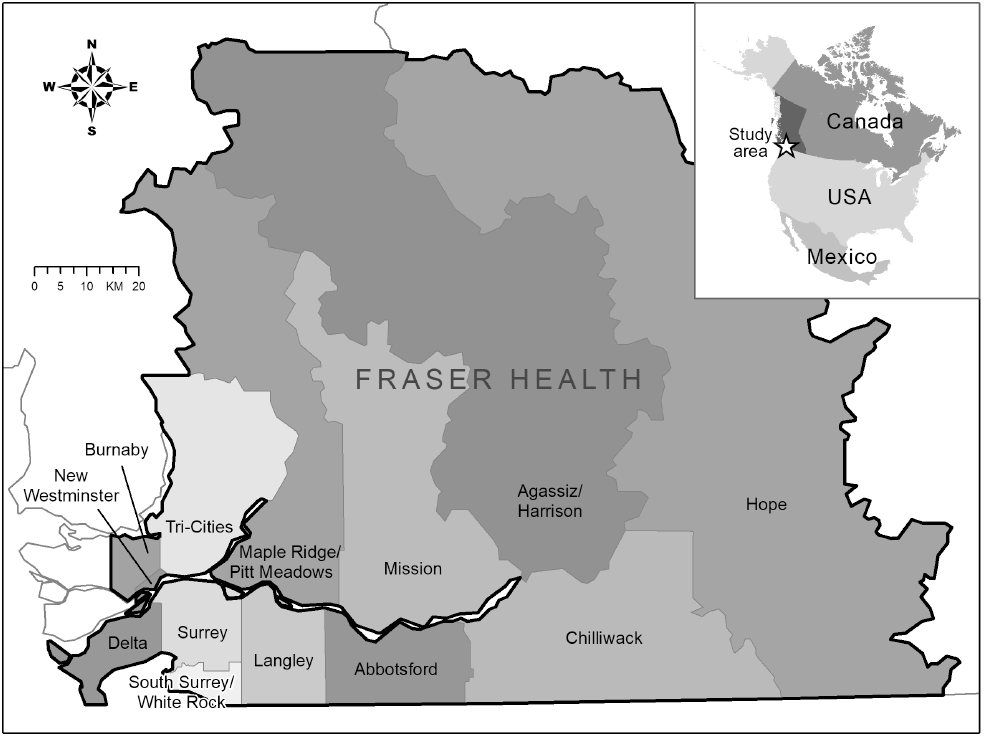
Map of the local health areas in Fraser Health, British Columbia (BC), Canada. The population size of each region is given in Table S3 of Supplementary material A.5.

### 2.2 Data

Human population move between regions for many reasons including work, leisure, family visits, health reasons, e.t.c. The main goal of this work is to develop a mathematical modeling framework for studying and understanding the effect of mobility on the spread of COVID-19 within and between regions. We consider the period from July 1, 2020 to January 27, 2021, inclusive. Although, movement restriction was imposed in Fraser Health during some part of this period, we used the mobility data collected through mobile device counts as a proxy for quantifying movements between the regions and the contact rate within each region.

We quantify mobility between the regions using two approaches. The first approach uses the physical distances between the regions, based on population weighted centroid (left panel of Figure 3) and the formula in (2.5) to calculate the probability that an individual moving in region *j*, who came from one of the 13 regions, originated from region *i* (*π*_*ji*_). The premise of using physical distance between regions is based on the concept of geographic distance decay, where spatial and social interactions decrease as the distance between regions increases [55]. The computed probabilities are presented in the right panel of Figure 3. The diagonal entries of this matrix represents the probability that an individual moving in a region is a resident of that region. It is important to note that the probability matrix is not symmetric, even though the distance matrix (left panel of Figure 3) is symmetric. In addition, each row of the probability matrix sums to 1. The second approach used to construct the probability matrix (*π*) is based on mobile device counts and uses Telus mobility (TELUS) data.

**Figure 3:**
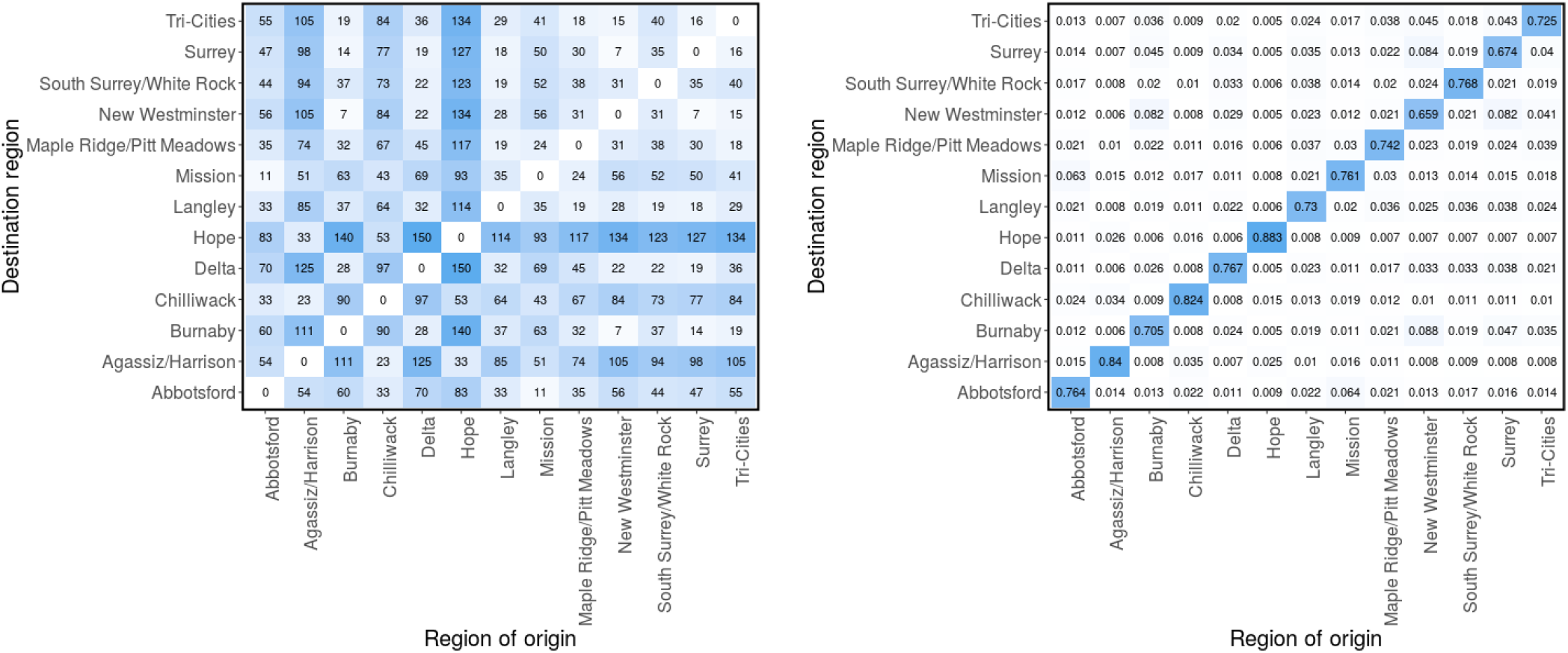
Distance matrices. Physical distances (in km) between the local health areas (LHAs) based on population-weighted centroid (left) [55] and the probability matrix (π) computed using (2.5) (right). π_ji_ is the probability that an individual who came from one of the 13 regions to region j, originated from region i.

TELUS is a Canadian national telecommunications company that has network coverage in 99% of the populated areas of Canada. TELUS Insights provides anonymized geo-intelligence data, which reflects population location and mass movement patterns based on information about locations and population movement of TELUS mobile device users [57]. These data have helped answer a range of questions around location and public mobility patterns within Canada, including in infrastructure planning, health services, roads, and transit routes. As TELUS subscribers use their mobile devices, they connect to various cellular towers for telecommunication services. Mobility data is determined from users who have actively turned on their ‘Location History’ setting in correspondence with their mobile phones global positioning system (GPS). This network data provides insights into movement patterns and trends across Canada. To provide a layer of privacy, all the mobility data provided by TELUS are de-identified, aggregated into large data pools, rounded-up to the nearest 10 counts and all results are extrapolated to represent the entire population of a given region. This ensures that the data cannot be traced back to individual TELUS subscribers. The results of the TELUS application programming interface (API) implementation, which provide the numbers of mobile devices moving within and between geographical locations of interest and the neighbourhood that a mobile device resides in, depend on cellular tower locations at the time of the analysis.

We generate the mobility data for each region using a one-day bucket size and 120-minute minimum dwell time. We filtered for “non-residents”, “moving residents” and “residents”, which represent, respectively, the daily number of mobile devices residing in an LHA and spending at least two hours in another LHA (movement between regions), the daily number of mobile devices residing in an LHA and spends over two hours outside their census track within the LHA (movement within a region), and the total number of mobile devices residing in an LHA. To construct the weekly mobility matrices, we consider the “non-residents” and “moving residents” data. For each region and for a specified time interval (weekly), we compute the number of mobile devices from the other 12 regions that visited the region and stayed there for at least 2 hours during the visit. This gives us the mobile device count for movement into the region (off-diagonal entries). For movement within the regions (diagonal entries), we used the “moving residents” data, from which we computed the number of mobile devices registered to a region and moving within the region. These information are used to construct a mobility matrix of device counts within the specified time interval for each region. We normalize each row of the matrix with the total number of devices in the row. This way, the *i*^th^ element of the *j*^th^ row represents the fraction of mobile devices that came into the *j*^th^ region (from the 13 regions) that originated from the *i*^th^ region. These fractions can also be interpreted as the probability that an individual moving within the *j*^th^ region (whom originated from one of the 13 regions) is from the *i*^th^ region (*π*_*ji*_). Using this approach we compute the probability/mobility matrices for each week from July 1, 2020 to January 27, 2021. The computed matrices for week 1 (July 1-7, 2020) and 30 (January 21-27, 2021) are shown in Figure 4, while the matrices for the remaining weeks are presented in Figures (S2 - S6) of Supplementary material A.2. The distance matrix (right panel of Figure 3) and the constructed mobility matrices are used to describe the interaction between individuals from different regions. We considered two main scenarios in our Bayesian inference based on the distance and mobility matrices and investigated whether the mobility data is more informative, in terms of disease transmission than the distances between the regions.

**Figure 4:**
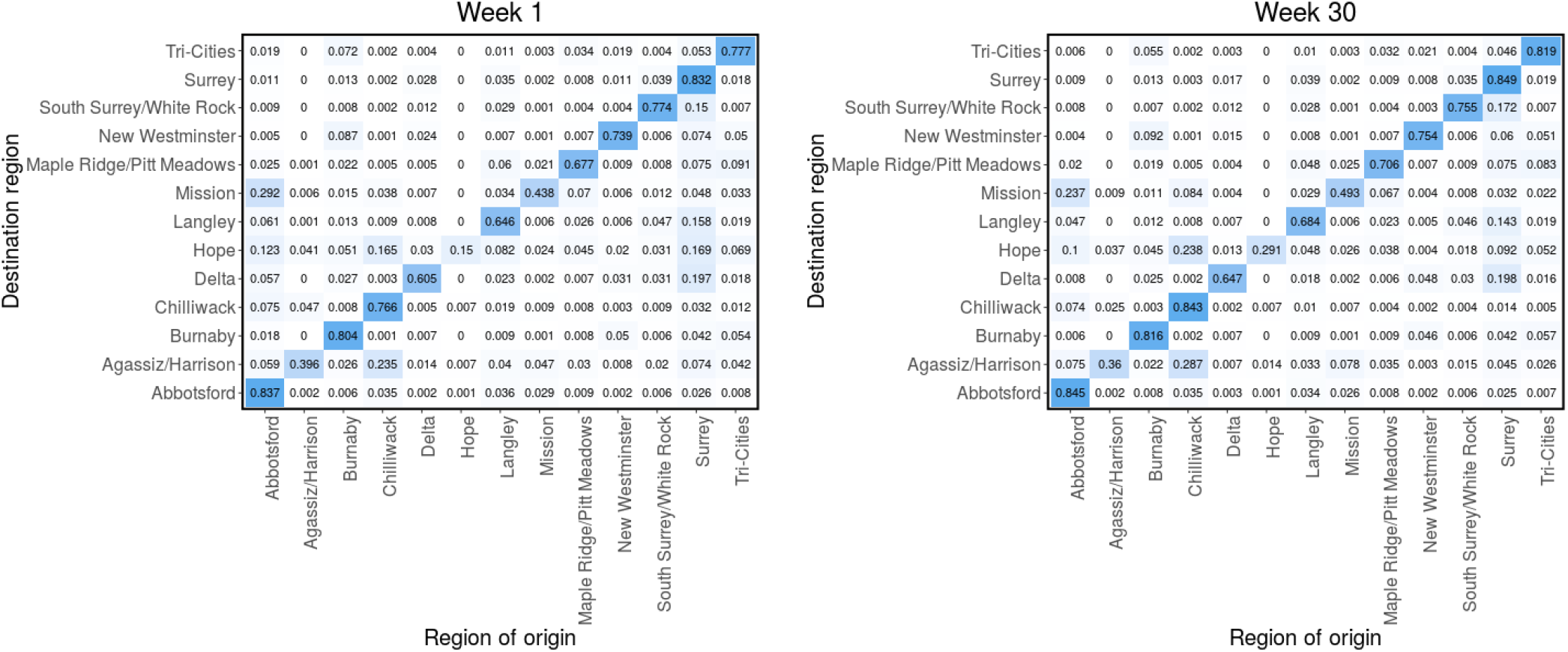
Mobility matrices. Probability matrix (π) computed from the Telus mobility data for week 1 (left) and week 30 (right), corresponding to July 1-7, 2020 and January 21-27, 2021, respectively. π_ji_ is the probability that an individual who came from one of the 13 LHAs to region j, originated from region i. Mobility matrices for the remaining weeks are presented in Figures (S2 - S6) of Supplementary material A.2.

We also used the Telus mobility data to compute the weekly mobility rate for each region. To compute these rates, we sum the daily device count in each region for “non-residents” and “moving resident”, and divide it by the sum of the “residents” and “non-residents” device count for our entire study period. This gives us the proportion of mobile devices moving in each region with respect to the total number of devices in the region during our entire study period. For each week in our study period, we sum the computed proportion of mobile devices and divide by 7 to get the weekly average proportion of mobile devices moving in each of the regions, as shown in Figure 5. These mobility rates are used as proxy for the contact rates in the regions and are represented by *m*_*j*_(*t*) is the disease transmission rate (*β*_*j*_(*t*)) defined in (2.2). We observe from Figure 5 that there is a sharp decline in mobility rate around the first week of September 2020 in most of the regions. Similarly, there is another decline in mobility rate around the first week on November. This decline is associated with the implementation of public health measures in BC.

**Figure 5:**
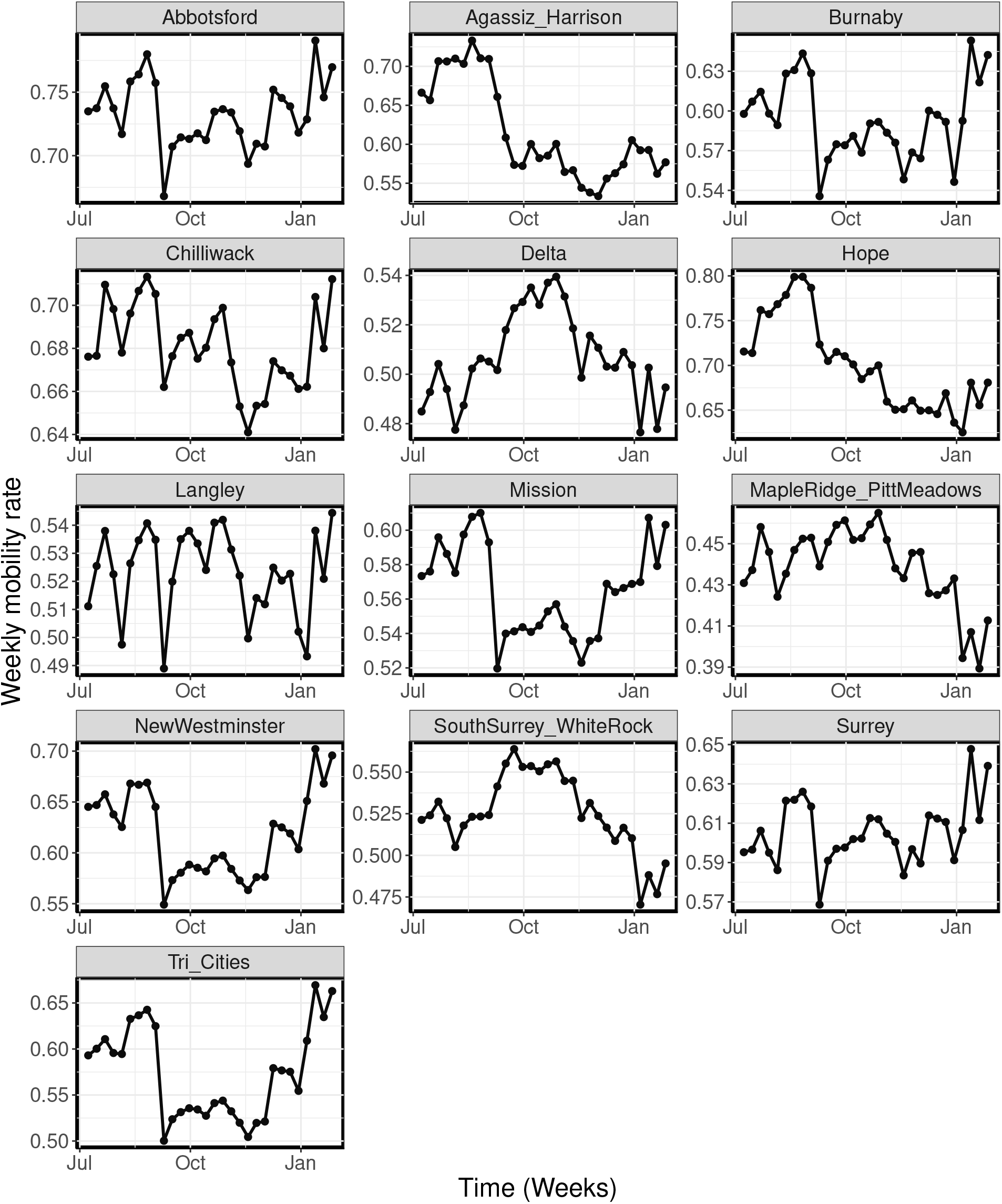
Weekly mobility rates. Weekly time-series mobility rates for each region from July 2020 - January 2021, computed from the Telus mobility data.

We calibrate our model to the weekly reported cases of COVID-19 in the thirteen local health areas of Fraser Health, BC, obtained from the British Columbia Centre for Disease Control (BCCDC). We extracted these data from a line list generated by BCCDC Public Health Reporting Data Warehouse (PHRDW), based on symptom onset date or reported date where symptoms onset date is not available. The collected case data spans the period from July 2020 to January 2021, inclusive, and was incorporated into the model likelihood based on the computed disease incidence as shown in (2.6). The collected weekly reported cases of COVID-19 for the 13 regions are shown in Figure S1 of Supplementary material A.1. Similar to [35], our model incidence is computed as the number of individuals in the pre-symptomatic population (*P*), transitioning to the infectious compartment (*I*_1_).

### 2.3 Bayesian inference

Our hybrid gravity-metapopulation model (2.1) is fitted to the COVID-19 cases in all the thirteen regions simultaneous using a Bayesian hierarchical modeling framework and the RStan package in R version 3.6.3 [54]. For the *j*^th^ region, we construct the likelihood as

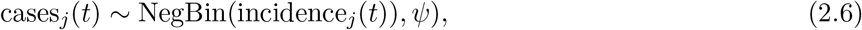

where NegBin(·) is the negative binomial distribution, cases_*j*_(*t*) and incidence_*j*_(*t*) are the weekly reported cases of COVID-19 and the incidence computed from the model (2.1), respectively, for region *j*. The

Bayesian inference framework gives us the flexibility to incorporate our prior knowledge into the model parameters and the ability to evaluate probabilistic statements of the data based on the model. In addition, the hierarchical modelling framework allows us to construct the posterior distribution for the population mean and variance of the model parameters and those of the individual parameters for each region, which are conditioned on the population mean and variance. Uninformative priors were implemented in the Bayesian inference framework.

We incorporate the time-series mobility data (Figure 5) into our modeling framework using an exponential scaling approach for the disease transmission rate. The disease transmission rate is given by (2.2), where *c*_0*j*_ *∼ 𝒩*_+_(0, 1) is the scaling parameter for the baseline transmission rate for the *j*^th^ region (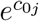 is the baseline transmission rate) and *c*_1_ ∼ 𝒩_+_(0, 1) is the scaling parameter used to remove biases from the time-series mobility data (*m*_*j*_(*t*)). Here, 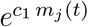 models the time-varying effect of *m*_*j*_(*t*) on the disease transmission rate (*β*_*j*_) for region *j*. The time-dependent piece-wise constant parameter *g*(*t*) is used to account for other factors that affect disease transmission, which are not explicitly accounted in the model. This parameter vector is estimated every four weeks (except for the last interval which has 2 weeks). We also estimated the total prevalence of COVID-19 in all the 13 regions at the beginning of our study period. Similar to [4, 35], when building our Bayesian inference modeling framework, we simulated the incidence for the metapopulation model (2.1) using known parameters values and then tested the ability of our framework to recover the values. We inspect the resulting posterior distribution for biases and their coverage of the true parameters.

Throughout this paper, we used the Variational Bayes (VB) method with the *meanfield* algorithm implemented in RStan [56, 69] for our inferences, from which we estimate the total initial prevalence in all the 13 region and a parameter (*θ*) used to quantify the effect of mobility on disease transmission in the regions (see the formulation in (2.4)). In addition, we used Bayesian hierarchical modeling framework to construct the posterior distribution for the mean and variance hyper-parameters for *c*_0*j*_, which are then used as prior distributions for the estimates of these parameters for each region, *c*_0*j*_ for *j* = 1, …, 13. Similarly, we construct the posterior distributions for the population mean and variance for the parameter *g*(*t*), which are then used as prior mean and variance, respectively for the estimated values of *g*(*t*) for each interval. We estimated a fixed value of the parameter *g*(*t*) for every four weeks, starting from the beginning of our study period, and for the last two weeks. Thereby making it a time-dependent and piece-wise parameter. To ensure that the estimated parameters are identifiable and that the estimated values of *g*(*t*) from the second interval onward are relative to that of the first interval, for each region, we rescaled the time-series mobility data by using the first week’s mobility rate as a reference for the remaining rates. This was done by subtracting the mobility rate for the first week from the weekly rates for the entire study period. This way the rescaled mobility rate for the first week is 0, while those for the remaining weeks are centered around 0. Similarly, *g*(*t*) for the first four weeks (first sub-interval) was set to 0. The remaining parameters of the model are fixed and are as presented in Table 1.

We considered two main scenarios in our model calibration: one with a fixed distance matrix (computed from the distances between the regions, see Figure 3) and another with weekly mobility matrices (computed from Telus mobility data, see Figure 4 and Supplementary material A.2). These two matrices are used to quantify mobility between the 13 regions. Performing inference based on these two scenarios enabled us to understand the effect of mobility on the posterior predictive distributions of the model and to determine which of the two mobility quantifiers best recreates the observed case data. It would also help us to identify, which of the two approaches provides more information on human mobility in terms of disease transmission. The two scenarios were ranked by comparing their leave-one-out predictions and standard errors, computed using the leave-one-out cross-validation (LOO) method [43, 17, 60], and using the widely applicable information criterion (WAIC) method [61, 16]. We compared the Variational Bayes method to the adaptive Hamiltonian Monte Carlo method No-U-Turn sampling. The results from both methods are found to produce comparable estimates of the posterior distribution with significant reduction in total computation time [33]. For the case of a fixed distance matrix, the mean and/or median ELBO usually converges in 5, 000 *−* 6, 000 iterations of the stochastic gradient ascent algorithm, while it converge in 11, 000 *−* 12, 000 iteration for the weekly mobility matrices case (see [34, 69, 24] for more information about ELBO in the variational Bayes method).

## 3 Results

We considered two main scenarios when fitting our model to the weekly reported cases of COVID-19 (see Methods). Results for the two scenarios, for selected regions (Agassiz/Harrison, New Westminster, Maple Ridge/Pitt Meadows and Surrey), are presented in Figure 6. We selected these regions based on their population sizes and geographical locations, to show the diversity in reported cases and population sizes in the regions considered, and the model’s ability in predicting cases irrespective of these factors. The results for the remaining regions are presented in Figures S7 and S8 of Supplementary material A.3.

**Figure 6:**
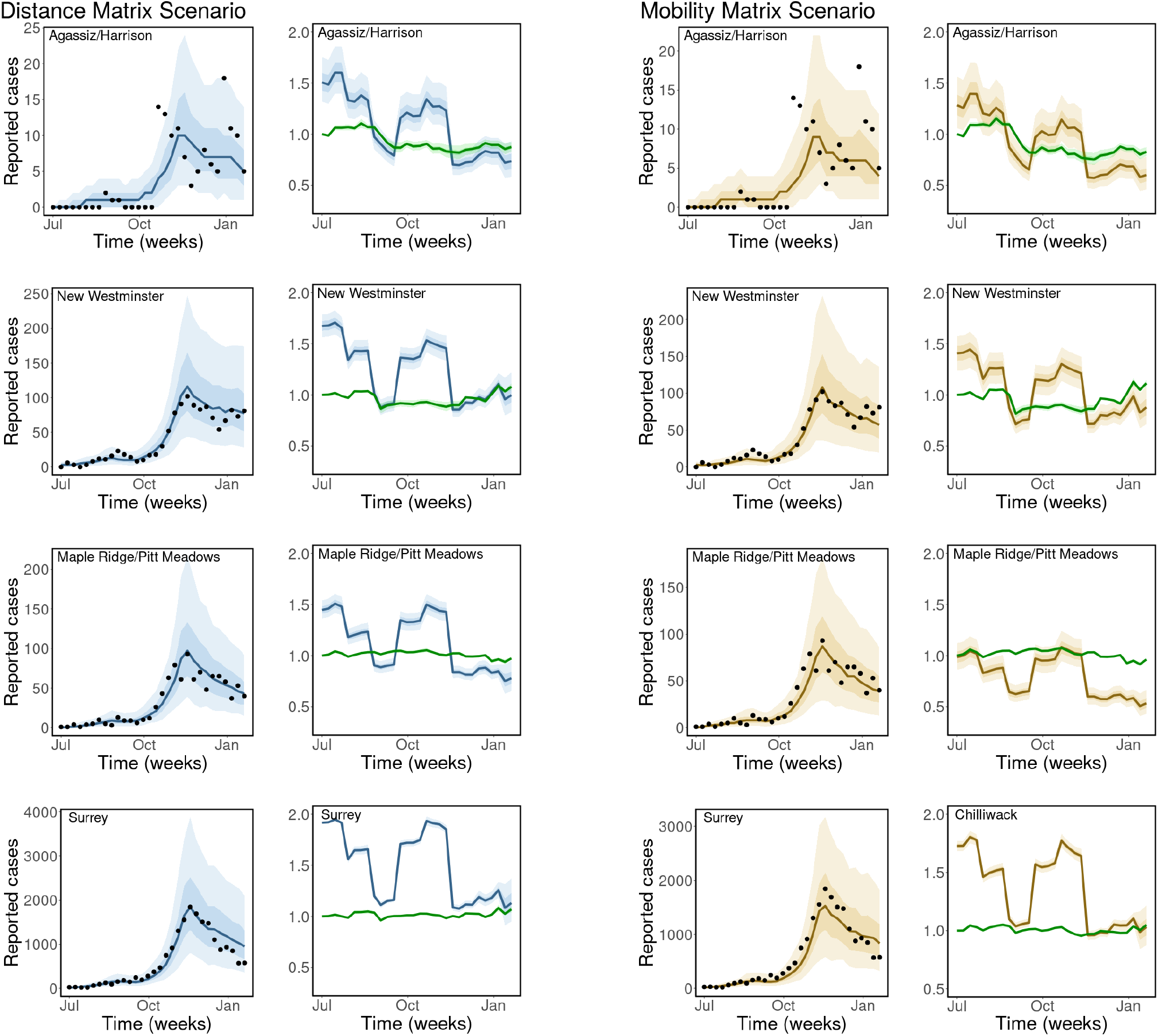
Observed and estimated COVID-19 cases. Weekly reported cases of COVID-19 and model prediction (columns 1 and 3). Disease transmission rate, β_j_(t) and the contribution of mobility to disease transmission, 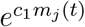 (green curves in columns 2 and 4). Model types: fixed distance matrix (blue) and weekly mobility matrices (gold). Black dots are the weekly reported cases of COVID-19, the solid lines are the mean estimates of cases/parameters, the darker bands are the 50% CrI, while the lighter bands are the 90% CrI. Similar results for the remaining regions are presented in Figures S7 and S8 of Supplementary material A.3.

For each model scenario, we present the posterior predictions of the weekly cases of COVID-19 in each region (columns 1 & 3 of Figure 6). We compute the time-dependent disease transmission rate, *β*_*j*_(*t*), using the estimated parameters and the formula in (2.2). These rates are presented in blue for the fixed distance matrix scenario (column 2 Figure 6) and in gold for the weekly mobility matrices scenario (column 4 of Figure 6), together with the contribution of mobility to the transmission rate, 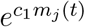 (green) with 50% credible interval (CrI) (darker bands) and 90% CrI (lighter bands). We observe from these results that our model is able to capture the trends and reported cases of COVID-19 in each of the regions with a high degree of accuracy for both model scenarios. In addition, we see that there are significant changes in the computed disease transmission rate over time, which has a similar trend for all the regions. Even though there are no much changes in the time-series mobility data, its effect on the disease transmission rates is still noticeable in the plot for each region.

The mean estimate for the initial total prevalence in the 13 regions is 47.61 (90% CrI: 44.82 - 50.31) for the distance matrix scenario and 50.19 (90% CrI: 47.37 - 53.04) for the weekly mobility matrix scenario. The mean estimate of the parameter used to quantify the effect of mobility on disease transmission in the regions (*θ*) for the distance matrix scenario is given by 0.53 (90% CrI: 0.44-0.60) and 0.90 (90% CrI: 0.72-0.98) for the scenario with weekly mobility matrices. This implies that movement between the regions contribute to a mean fraction of 0.53 and 0.90 of the total reported cases of COVID-19 in the regions for the distance and mobility matrix scenarios, respectively. The scaling parameter used to remove biases in the time-series mobility data (*c*_1_) was estimated as 1.51 (90% CrI: 0.90 - 2.10) for the distance matrix and 2.11 (90% CrI: 1.52 - 2.69) for the mobility matrix scenario.

We estimated the scaling parameters for the baseline disease transmission rate, *c*_0*j*_ for *j* = 1, …, 13, using Bayesian hierarchical modeling framework. These parameters are used to compute the baseline disease transmission rate for each region defined by 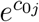 for *j* = 1, …, 13. The mean estimate for the hyper-mean and -variance are 0.45 (90% CrI: 0.35 - 0.54) and 0.18 (90% CrI: 0.12 - 0.26), respectively, for the distance matrix and 0.21 (90% CrI: 0.05 - 0.36) and 0.35 (90% CrI: 0.24 - 0.47), respectively, for the mobility matrix scenario. The mean estimate for *c*_0*j*_, for *j* = 1, …, 13 with 90% credible interval (CrI) are presented in Tables S1 (distance matrix) and S2 (mobility matrix) of Supplementary material A.4. The estimated distribution for the baseline disease transmission rates for the regions are presented in Figure 7. We observe from the results in this figure that the predicted distributions for the larger and more urbanized cities with dense population are similar for the distance and mobility matrix scenarios. These cities include Abbotsford, Burnaby, New Westminster, Surrey, and Tri-Cities. On the other hand, the predictions for the less densely populated smaller cities are relatively different for the two scenarios. In addition, the variance of the distributions for the smaller cities is larger than that of bigger cities with larger populations.

**Figure 7:**
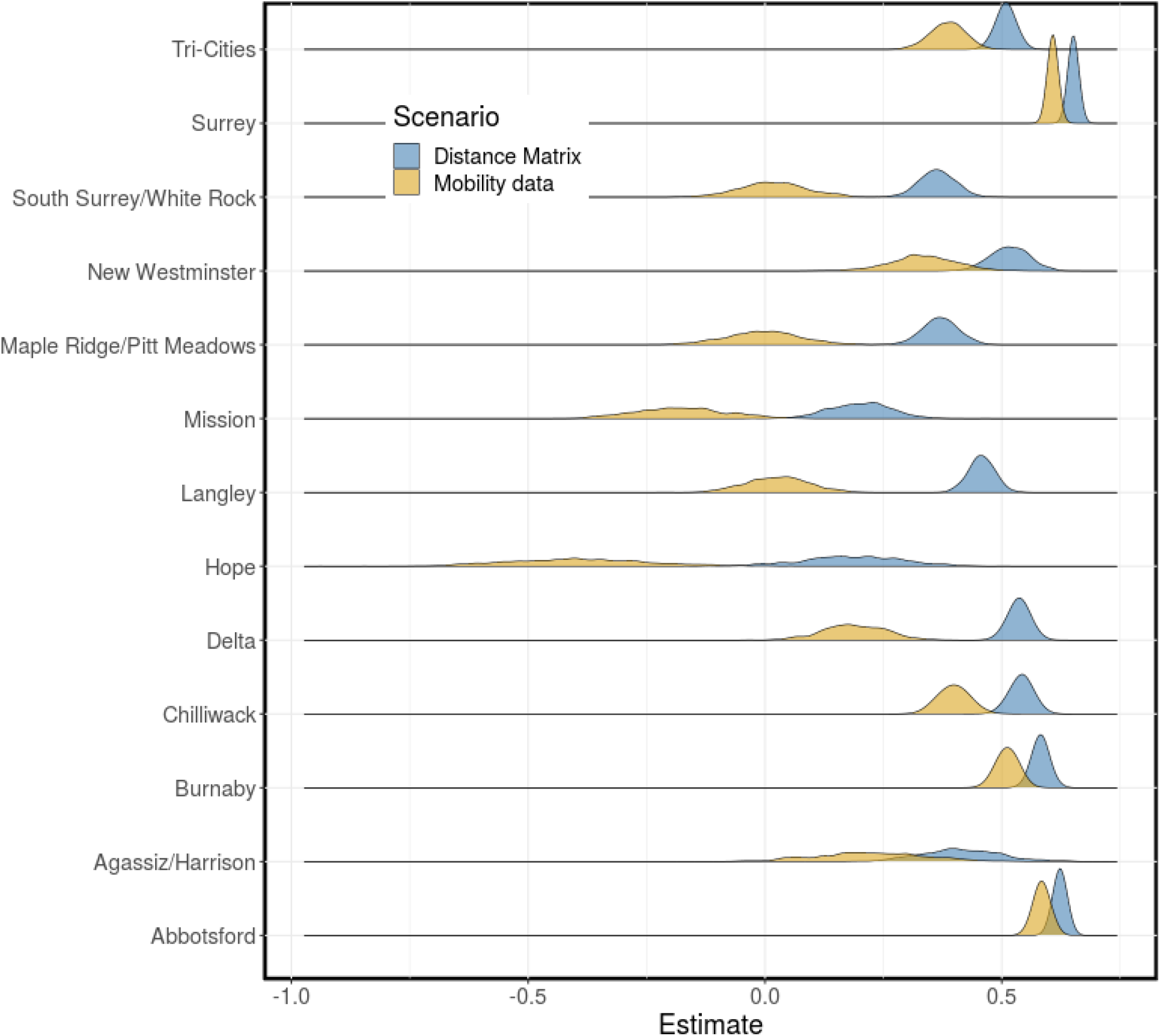
Baseline disease transmission rate. The distributions for the baseline disease trans-mission rate, 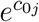, for each region computed using the estimated parameters c_0j_ for j = 1, …, 13 (see Tables S1 and S2 of Supplementary material A.4 for the estimates of c_0j_). Scenarios: fixed distance matrix (blue) and weekly mobility matrices (gold).

The time-dependent piece-wise parameter, *g*(*t*), was also estimated using a Bayesian hierarchical modeling framework with population mean and variance estimate with 90% credible interval given by -0.33 (−0.52, -0.14) and 0.30 (0.16, 0.47), respectively, for the distance matrix scenario, and -0.28 (−0.45, -0.10) and 0.32 (0.19, 0.50), respectively, for the weekly mobility matrix scenario. The mean estimates with 90% credible interval for the interval-specific parameters (*g*_2_ *− g*_8_) are given in Tables S1 (distance matrix scenario) and S2 (mobility matrix scenario) of Supplementary material A.4. It is important to emphasize that we have set *g*_1_ = 0 (week 1-4) to ensure that the model parameters are identifiable, and to estimate *g*_2_, …, *g*_8_ relative to *g*_1_. The distributions for the time-dependent effect of other factors that affect disease transmission, other than mobility, on the disease transmission rate *β*(*t*), are given in Figure 8. We observe that the constructed distributions for the two scenarios agree well.

**Figure 8:**
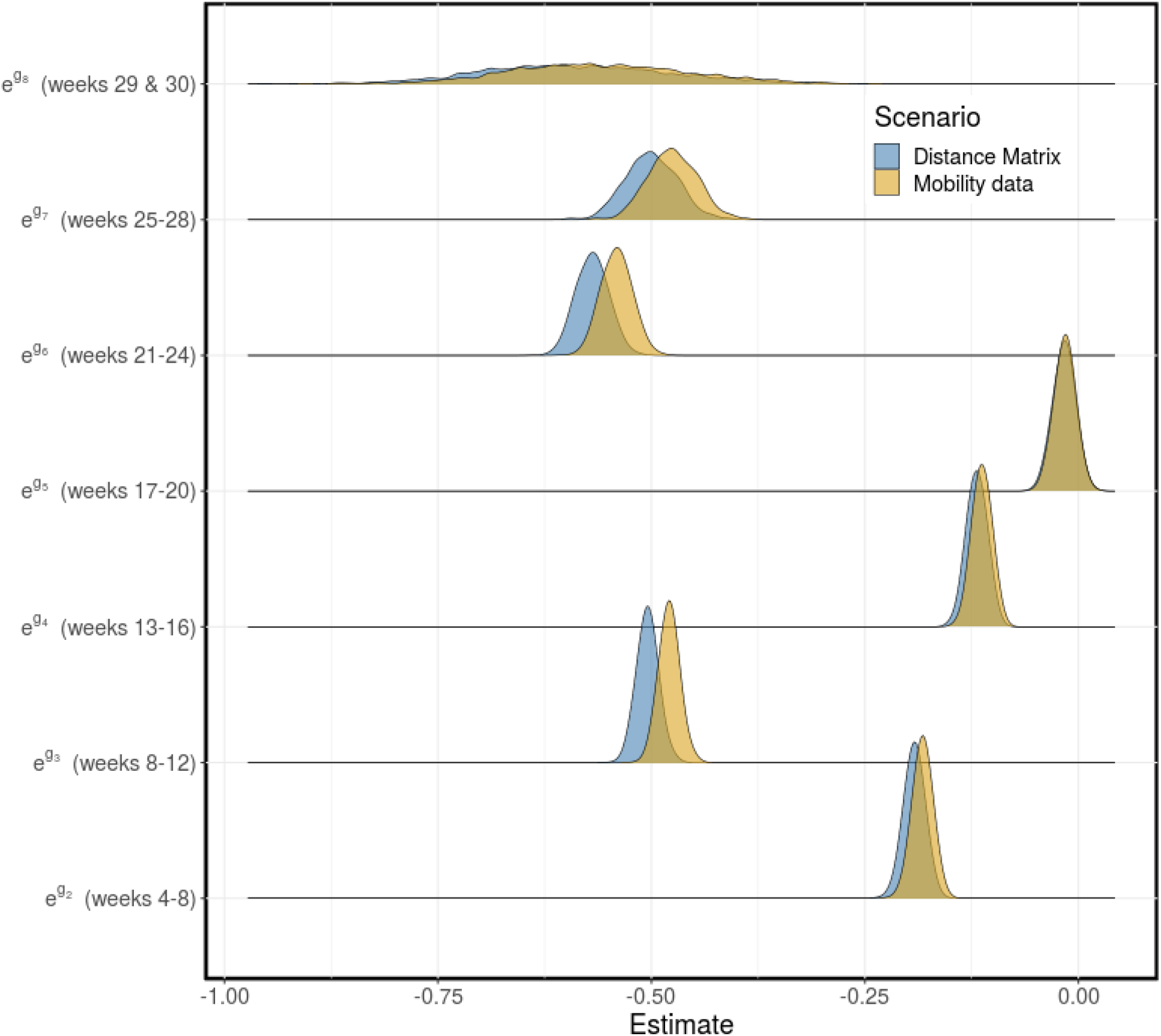
Contribution of other factors to disease transmission. The distributions for the contribution of other factors that affect disease transmission (e^g(t)^) to the transmission rate (β(t)), computed every four weeks and for the last two weeks. g_1_ = 0 (weeks 1-4), g_2_ (weeks 5-8), g_3_ (weeks 9-12), g_4_ (weeks 13-16), g_5_ (weeks 17-20), g_6_ (weeks 21-24), g_7_ (weeks 25-28), g_8_ (weeks 29 & 30). Scenarios: fixed distance matrix (blue) and weekly mobility matrices (gold). The estimated means with 90% credible interval for are presented in Tables S1 and S2 of Supplementary material A.4.

Lastly, we compare the estimated expected leave-one-out predictions and their standard errors, for the two model scenarios, computed using the leave-one-out cross-validation (LOO) method [43, 17, 60] and the widely applicable information criterion (WAIC) method [61, 16]. The comparison is summarized in Table 2, where the distance matrix scenario is ranked better than the mobility matrix scenario, in terms of their ability to capture the case data. Even though the distance matrix scenario captures the case data better than the weeekly mobility matrix scenario, the difference in the fits for the two approaches is not much.

**Table 2:**
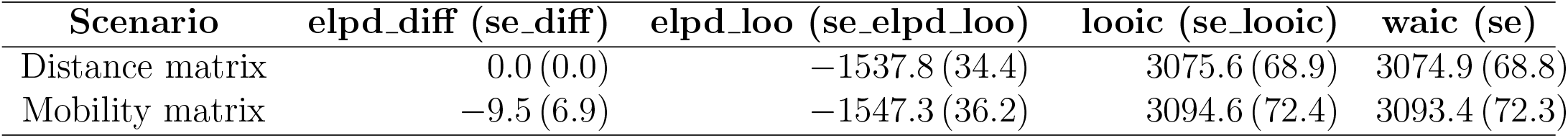
Model comparison using leave-one-out cross-validation (LOO) and the widely applicable or Watanabe-Akaike information criterion (WAIC). Model ranking (in descending order) is shown in the first column. The difference between the expected log pointwise predictive density (elpd) and its standard error (se elpd) for the best model and those of the remaining models are shown in the second column. In the third column, we have the Bayesian leave-one-out estimate of out-of-sample predictive fit (elpd loo) and its standard error (se elpd loo). Lastly, the computed Watanabe-Akaike information criterion (waic) for each model is shown in the fourth column.

| Scenario | elpd_diff (se_diff) | elpd_loo (se_elpd_loo) | looic (se_looic) | waic (se) |
| --- | --- | --- | --- | --- |
| Distance matrix | 0.0 (0.0) | -1537.8 (34.4) | 3075.6 (68.9) | 3074.9 (68.8) |
| Mobility matrix | -9.5 (6.9) | -1547.3 (36.2) | 3094.6 (72.4) | 3093.4 (72.3) |

## 4 Discussion

An important feature of our modeling framework includes the formulation of the disease transmission rate as an exponential function of a linear combination of factors that affect disease transmission. This formulation allows for explicit incorporation of factors into the transmission rate. In the example presented in this article, due to lack of adequate data, only time-series mobility data was incorporated explicitly into the disease transmission rate. The effect of other factors that affect disease transmission was accounted for using a time-dependent piece-wise parameter. We attempted to incorporate the effect of facemasks into the model but could not get adequate data for facemask usage in each region. In this case, the disease transmission rate was formulated as follows

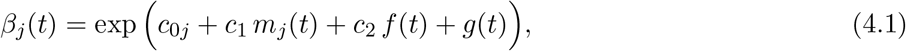

where *c*_0*j*_ for *j* = 1, …, 13 are region-specific scaling parameters used to compute the baseline disease transmission rate for each region (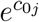 is the baseline transmission rate in region *j*). The parameters *c*_1_ and *c*_2_ are covariates for the mobility and facemask usage rates, respectively, and *g*(*t*) is a time-dependent piece-wise parameter that is used to incorporate the effect of other factors that affect disease transmission other than mobility and facemask. This formulation can also be extended to explicitly account for other factors that affect disease transmission in *β*_*j*_(*t*) based on data availability.

Our model was able to capture the trends and reported cases of COVID-19 cases in each region (see Figure 6, and Figures S7 and S8 of Supplementary material A.3). In addition, the results of the two model scenarios agree well, although, there is a slight different in the estimated time-dependent disease transmission rates and the contribution of mobility to disease transmission (columns 2 & 4) for some regions. There are significant changes in the computed disease transmission rate over time, which may be attributed to the intervention strategies implemented by the government during this period. Even though there are no much changes in the time-series mobility data, its effect on the disease transmission rate is still apparent for each region. The estimated total initial prevalence of COVID-19 in all the regions for the two scenarios agree well, as well as the estimates for the time-dependent piece-wise parameter (*g*(*t*)), used to incorporate the effect of other factors that affect disease transmission into the transmission rate (see Figure 8). However, the estimated effect of mobility on disease transmission is significantly different for the two scenarios. The mean estimate of this parameter was 0.53 (90% CrI: 0.44-0.60) for the distance matrix scenario and 0.90 (90% CrI: 0.72 - 0.98) for the weekly mobility matrix scenario. This can be interpreted as mobility contributing to 53% and 90% of the cases in the regions for the distance and mobility matrices scenarios, respectively. These results show that the weekly mobility data provides more information, in terms of disease transmission, than the distances between the regions. Note that the mobility referred to here is for both within and between the regions. To confirm that indeed the weekly mobility data provides more information, we considered a third scenario, where we used a fixed mobility matrix computed using the mobility data for the entire study from July 2020 to January 2021. For this scenario, we estimated the effect of mobility on disease transmission as 0.60 (90% CrI: 0.52 - 0.70) (see Supplementary material A.6 for more details). As expected, the fixed mobility matrix does not provide more information about disease transmission than the weekly mobility matrices, even though it does better than the distance matrix.

The constructed distributions for the baseline disease transmission rate for the two model scenarios are similar for some of the regions and significantly different for other regions. These distributions are similar for the larger and more urbanized cities with dense population (Abbotsford, Burnaby, New Westminster, Surrey and Tri-Cities) and significantly different for the less densely populated smaller regions (see Figure 7). The difference in the predicted distributions for these two groups of regions may be attributed to their population size and mobility in the regions. Lastly, we compared the results obtained from the two scenarios using the leave-one-out cross-validation (LOO) and the widely applicable information criterion (WAIC) methods. This comparison ranks the distance matrix results better than those of the weekly mobility matrices, although, the computed loo and waic for the two scenarios are very similar (see Table 2). We considered these two model scenarios in order to test the hypothesis of whether the time-dependent mobility matrices, computed from the mobile device data, provide more information about human mobility between the regions in terms of disease transmission than the distances between the regions. Based on our results, we conclude that even though the distance matrix provides a better fit to the data, the weekly mobility matrices have the ability to explain the variance in transmission between regions. The model for when the distance matrix is used is considered a *gravity model*, while the scenario where the weekly mobility matrices are used is referred to as a *metapopulation model*. Hence, our *hybrid gravity-metapopulation model*.

Overall, our modeling framework provides the ability to explicitly incorporate real data on factors the affect disease transmission into the disease transmission rate, and also allows independent assessment of the effect of these parameters on disease transmission in an epidemic. Furthermore, this framework allows us to quantify the effect of mobility on disease transmission in the regions. However, this work is not without limitations. We quantified the effect of mobility on disease transmission in the 13 LHAs of Fraser Health, BC, based on movements between these thirteen regions only. However, there are movement in and out of these regions to other parts of Canada, and even international. Another limitation of this work is that some regions in Fraser Health are closer to regions in other regional health areas in British Columbia, than they are to other regions in Fraser Health. For example, Burnaby is closer to Vancouver than it is to many of the LHAs in Fraser Health. As a result of this, the spread of COVID-19 in Burnaby may be influenced more by the number of cases in Vancouver than in other regions in Fraser Health, e.g. Hope, Chilliwack and Agassize/Harrision. In the example presented here, we explicitly incorporated only the time-series mobility data into the disease transmission rate and accounted for other factors that affect disease transmission through a piece-wise parameter. Interesting extensions of this work would be to incorporate the data for other factors that affect disease transmission explicitly into the model. This way, the effect of each factor on disease spread can easily be assessed. Another future work on this project is to extend the model to include vaccination and the variants of concern of COVID-19. Since mobility rate varies by age, another interesting extension of this work would be to stratify the population of each region by age. This way, in addition to assessing the impact of mobility on disease spread, it would also be possible to assess the contribution of each age group to disease spread.

## Data Availability

All data produced in the present work are contained in the manuscript.

## Acknowledgement

We acknowledge the support of TELUS and Michelle Spencer in making the mobile device data available.

## Data availability statement

All relevant data are within the paper and its Supplementary material. The Bayesian inference codes are available on GitHub at https://github.com/iyaniwura.

## A Supplementary material

### A.1 Weekly reported cases of COVID-19

We present the weekly reported cases of COVID-19 in the thirteen local health areas of Fraser Health, BC, Canada, from July 2020 to January 2021, inclusive. The data was extracted from a line list generated by BCCDC Public Health Reporting Data Warehouse (PHRDW), based on symptom onset date or reported date where symptoms onset date is not available.

**Figure S1:**
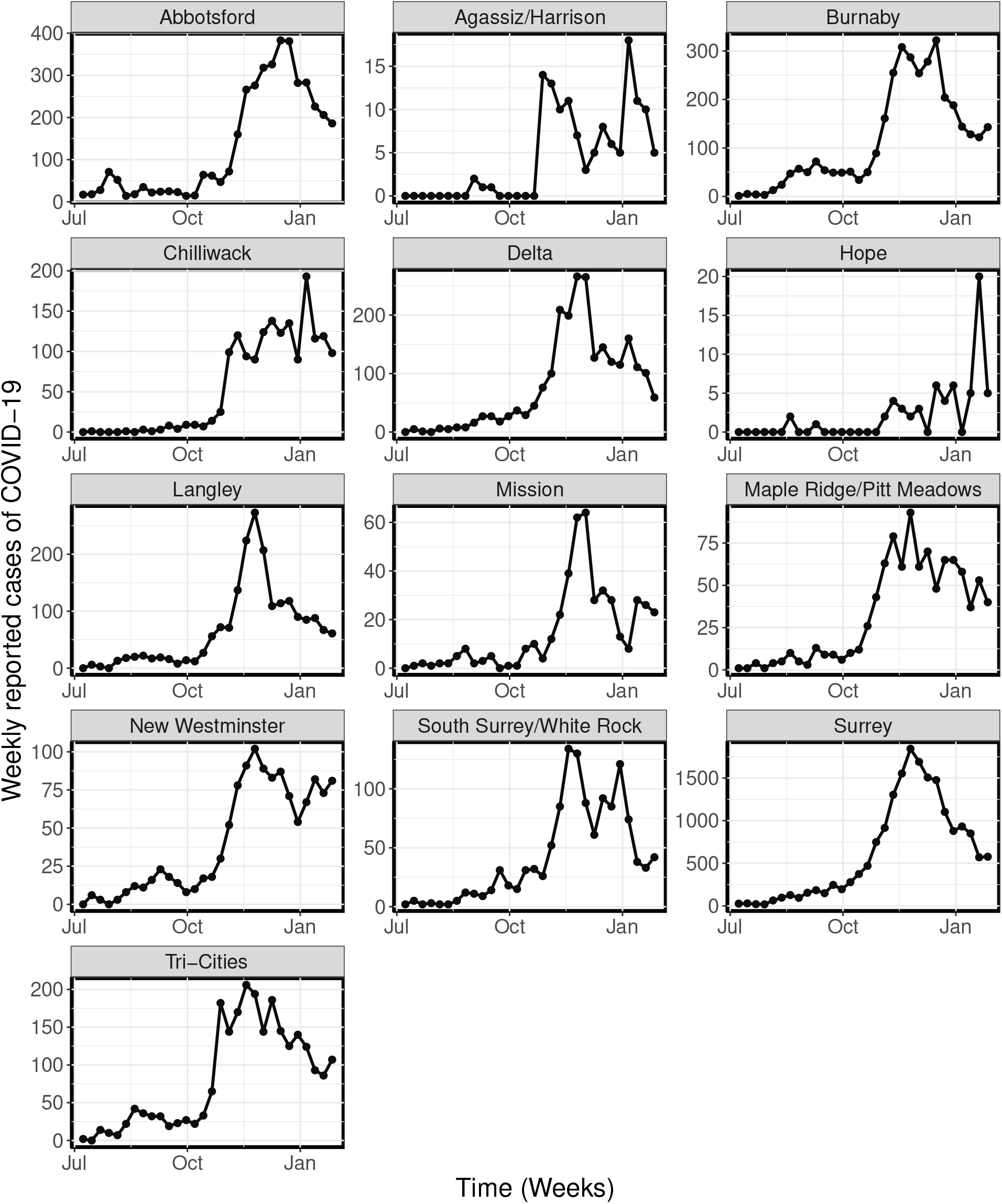
Reported cases of COVID-19. Weekly reported cases of COVID-19 for the 13 local health areas (LHA) of Fraser Health, British Columbia (BC), Canada, for the period from July 2020 to January 2021.

### A.2 Weekly mobility matrices

We present the weekly mobility matrices constructed from Telus mobility data for the period from July 2021 to January 2022.

**Figure S2:**
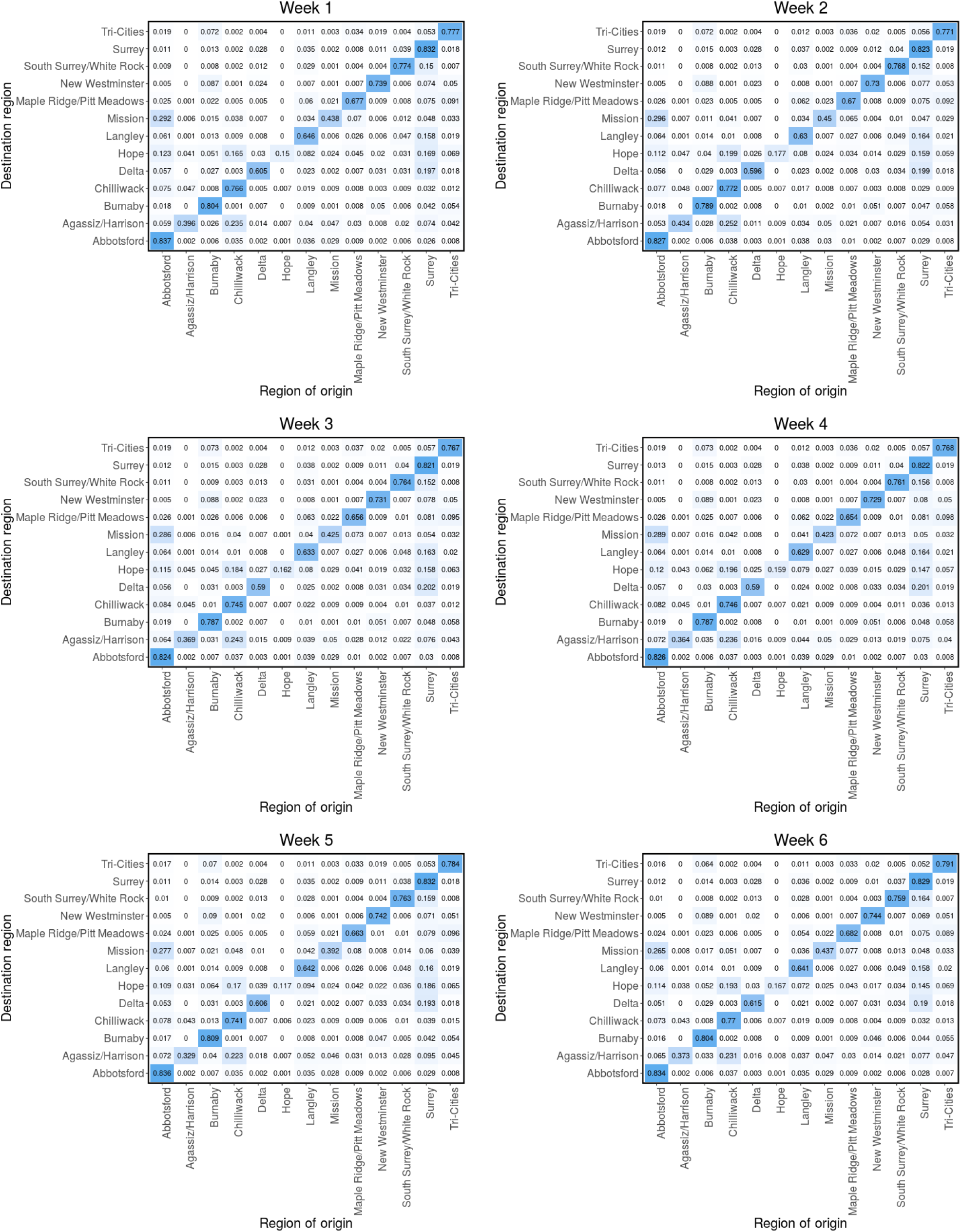
Mobility matrices. Probability matrix (π) computed from the Telus mobility data for week 1 - 6 (July 1, 2020 - Aug 12, 2020). π_ji_ is the probability that an individual who migrated from one of the 13 LHAs to region j, originated from region i.

**Figure S3:**
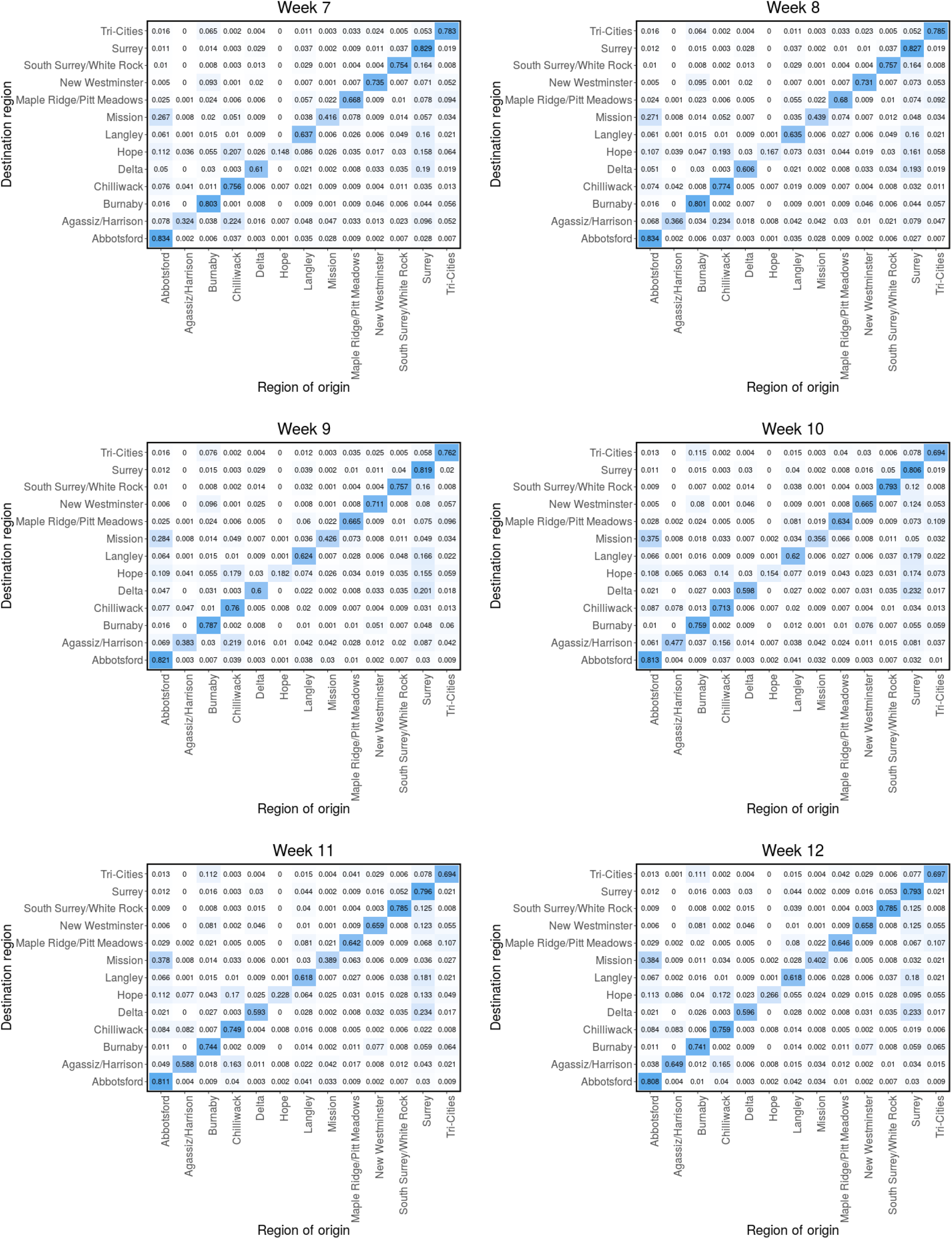
Mobility matrices. Probability matrix (π) computed from the Telus mobility data for week 1 - 6 (Aug 13, 2020 - Sept 23, 2020). π_ji_ is the probability that an individual who migrated from one of the 13 LHAs to region j, originated from region i.

**Figure S4:**
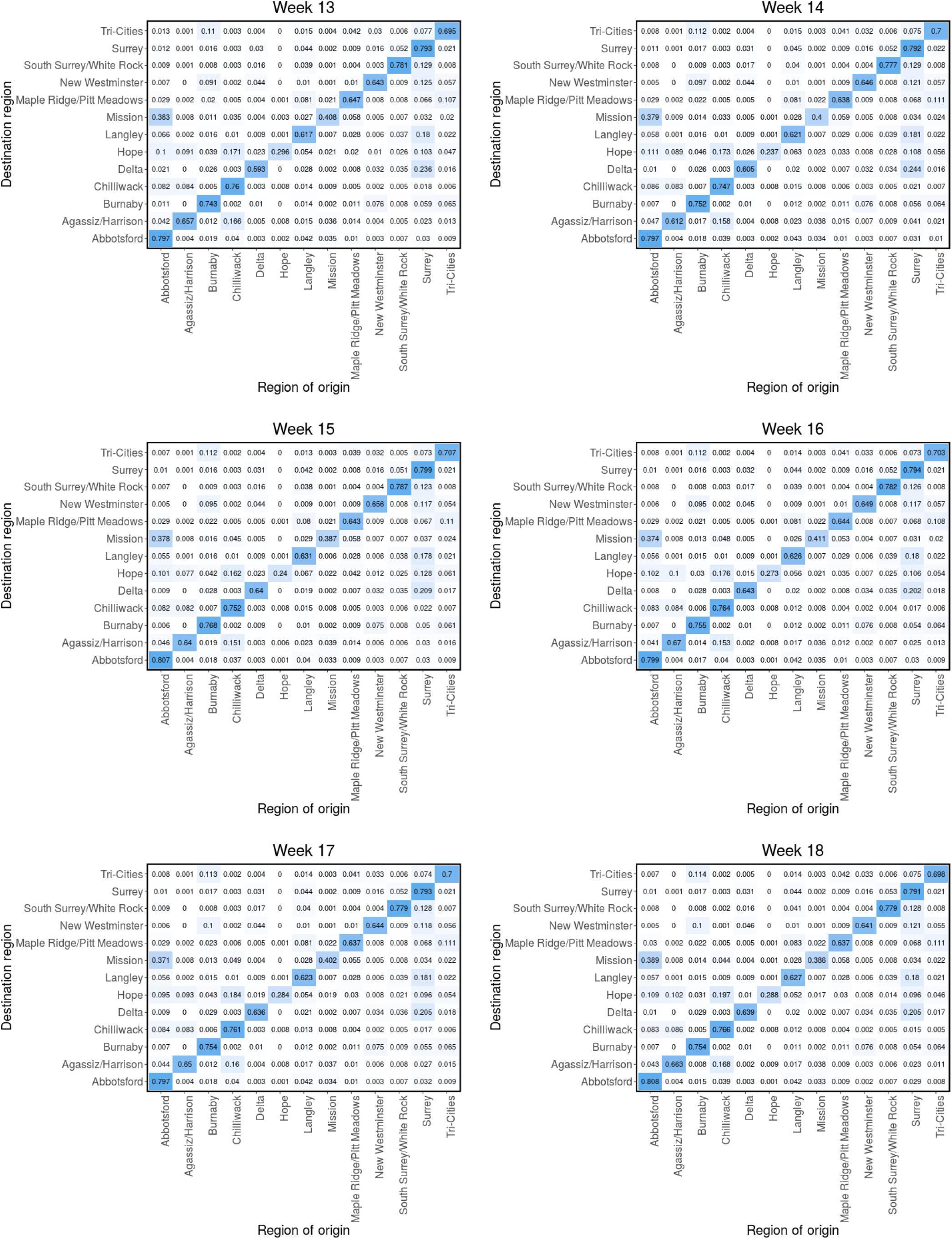
Mobility matrices. Probability matrix (π) computed from the Telus mobility data for week 1 - 6 (Sept 24, 2020 - Nov 4, 2020). π_ji_ is the probability that an individual who migrated from one of the 13 LHAs to region j, originated from region i.

**Figure S5:**
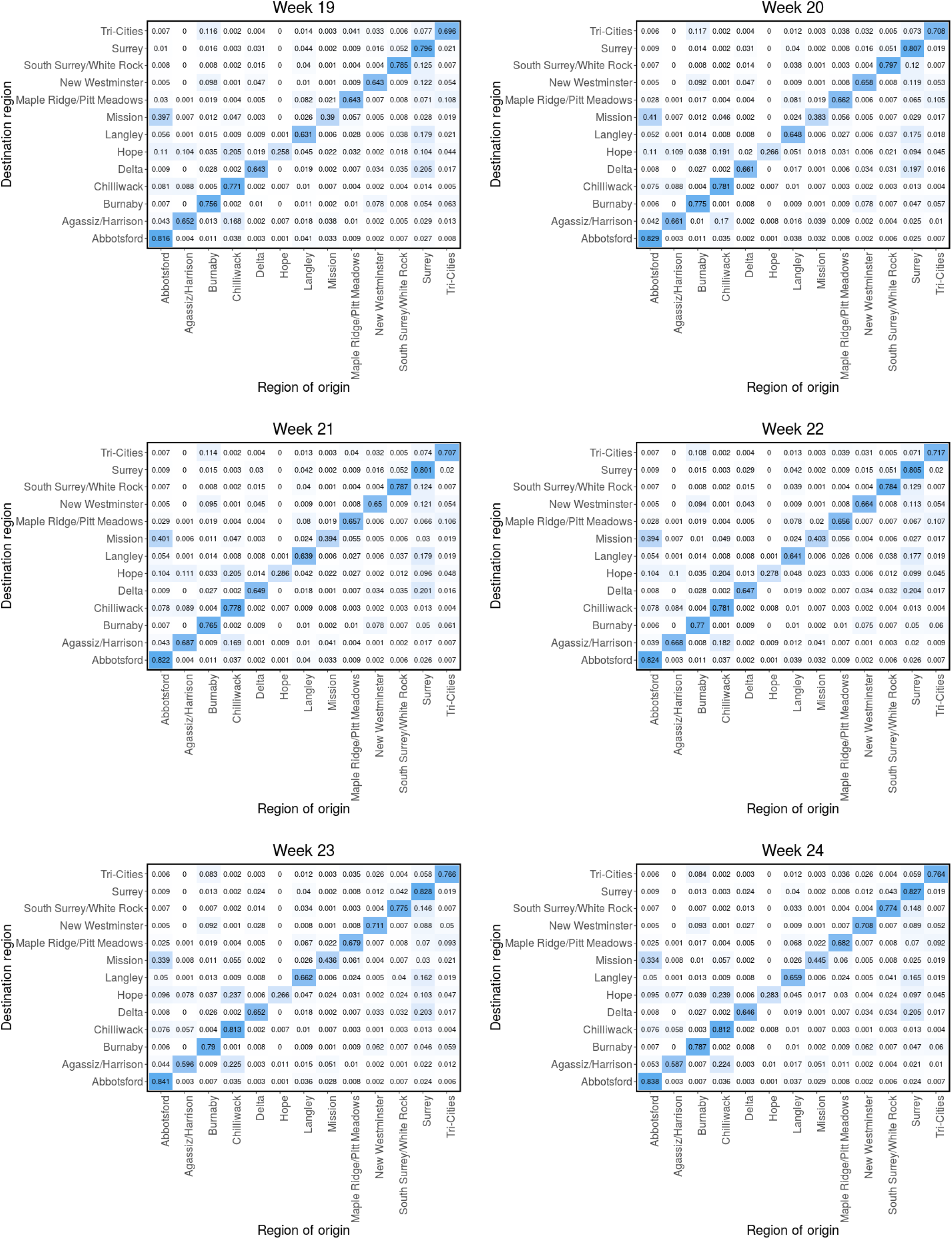
Mobility matrices. Probability matrix (π) computed from the Telus mobility data for week 1 - 6 (Nov 5, 2020 - Dec 16, 2020). π_ji_ is the probability that an individual who migrated from one of the 13 LHAs to region j, originated from region i.

**Figure S6:**
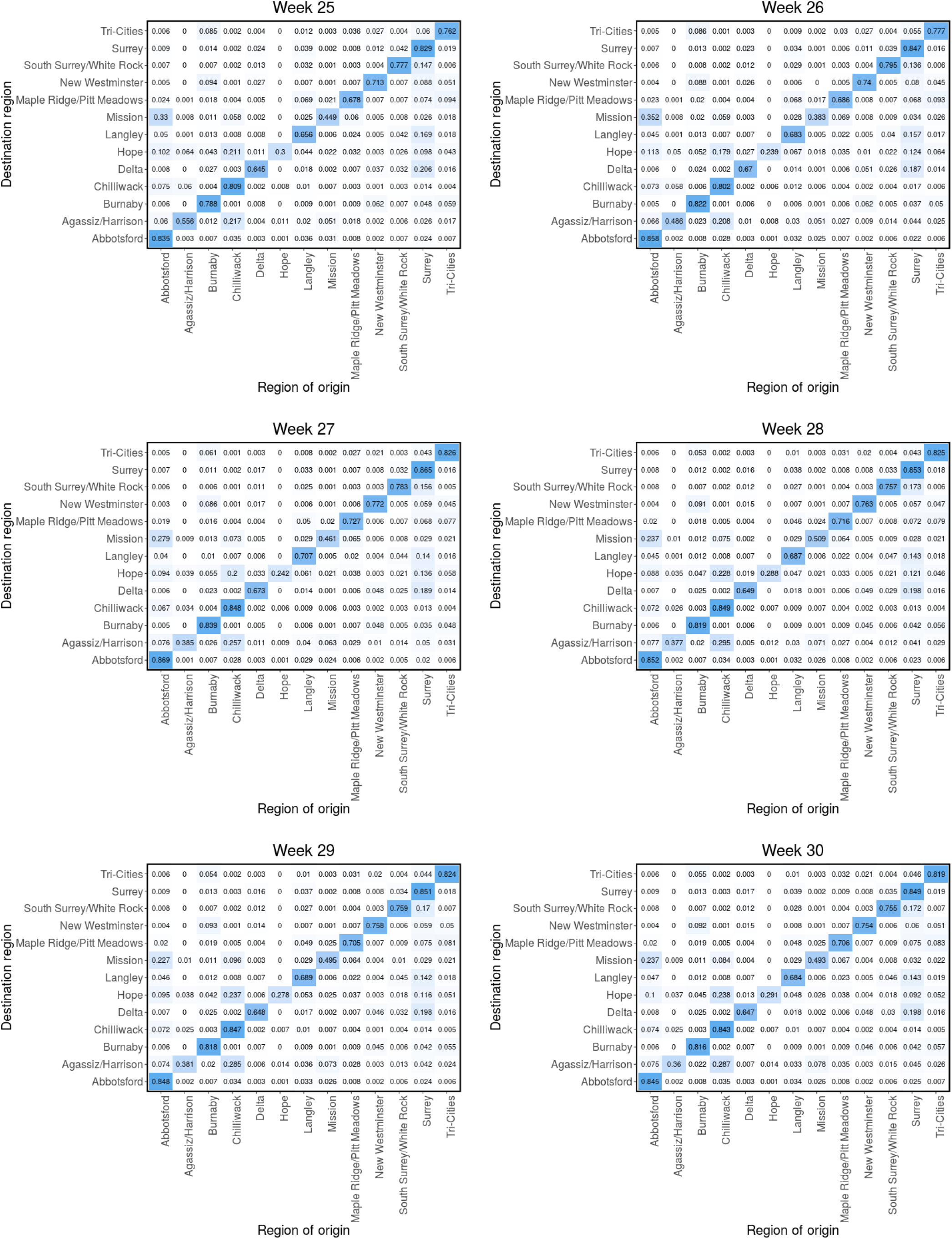
Mobility matrices. Probability matrix (π) computed from the Telus mobility data for week 1 - 6 (Dec 17, 2020 - Jan. 27, 2021). π_ji_ is the probability that an individual who migrated from one of the 13 LHAs to region j, originated from region i.

### A.3 Reported cases and model predictions

In this section, we present the model’s predicted cases and the estimated parameters for the remaining nine LHA whose results are not presented in the result section (§3).

**Figure S7:**
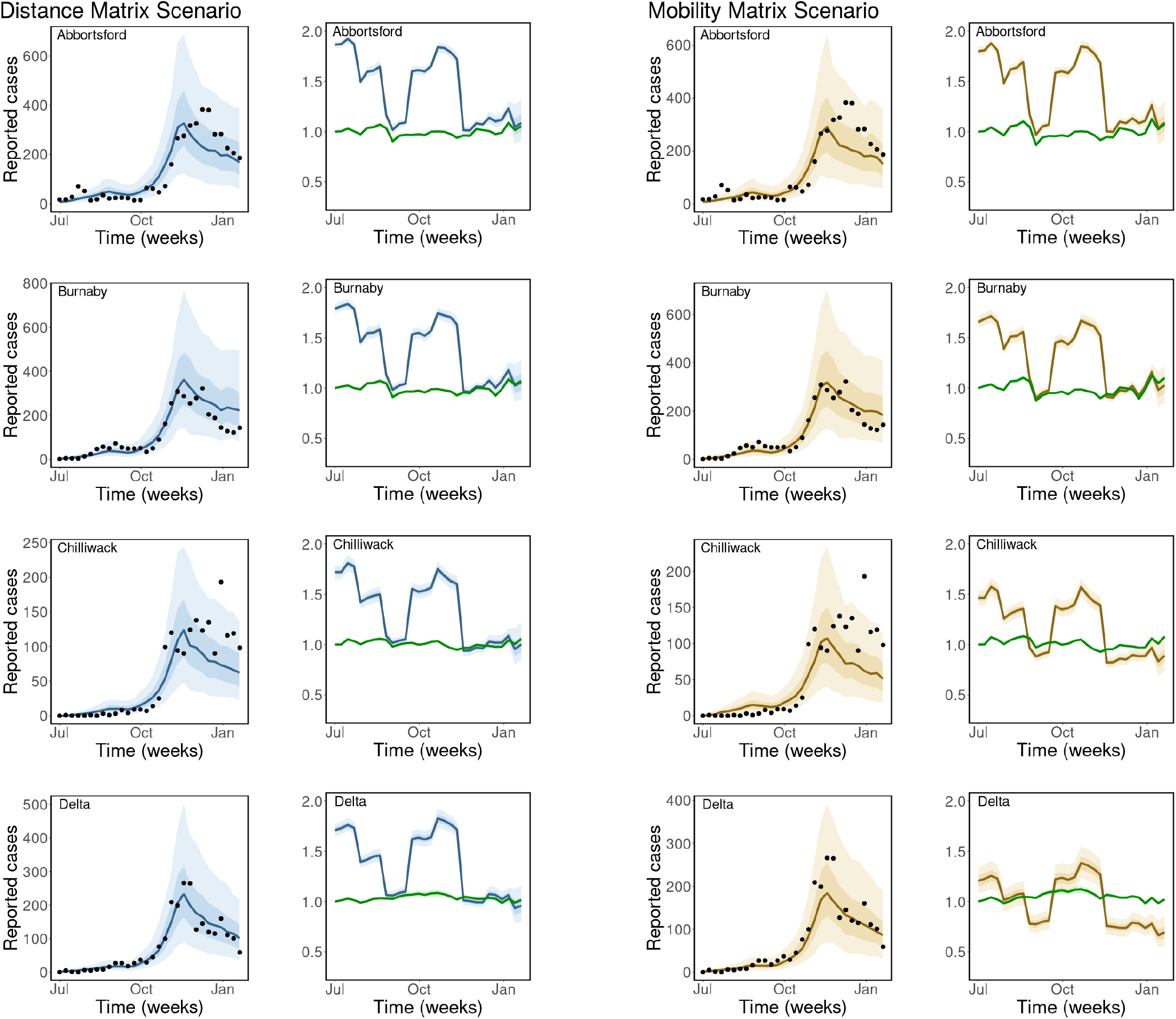
Observed and estimated COVID-19 cases. Weekly reported cases and model prediction (columns 1 and 3). Disease transmission rate, β_j_(t) and the contribution of mobility to disease transmission, 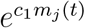 (columns 2 and 4). Scenarios: fixed distance matrix (blue) and weekly mobility matrices (gold). Black dots are the weekly reported cases of COVID-19, the solid lines are the mean estimates of cases/parameters, the darker bands are the 50% CrI, while the lighter bands are the 90% CrI.

**Figure S8:**
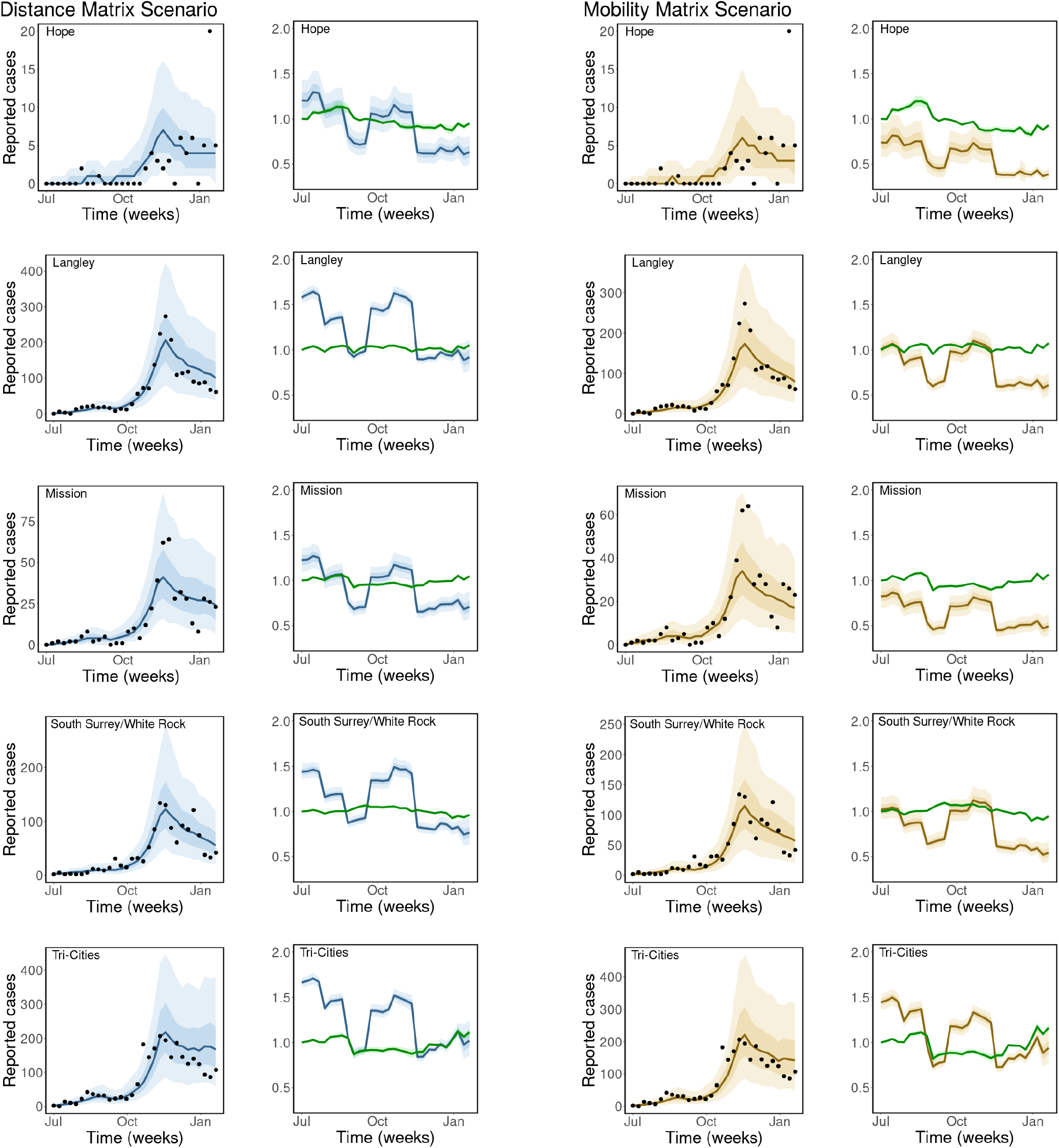
Observed and estimated COVID-19 cases. Weekly reported cases and model prediction (columns 1 and 3). Disease transmission rate, β_j_(t) and the contribution of mobility to disease transmission, 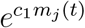 (columns 2 and 4). Scenarios: fixed distance matrix (blue) and weekly mobility matrices (gold). Black dots are the weekly reported cases of COVID-19, the solid lines are the mean estimates of cases/parameters, the darker bands are the 50% CrI, while the lighter bands are the 90% CrI.

### A.4 Table of estimated parameters

We present the estimated parameters for the distance matrix and weekly mobility matrices scenarios.

**Table S1:**
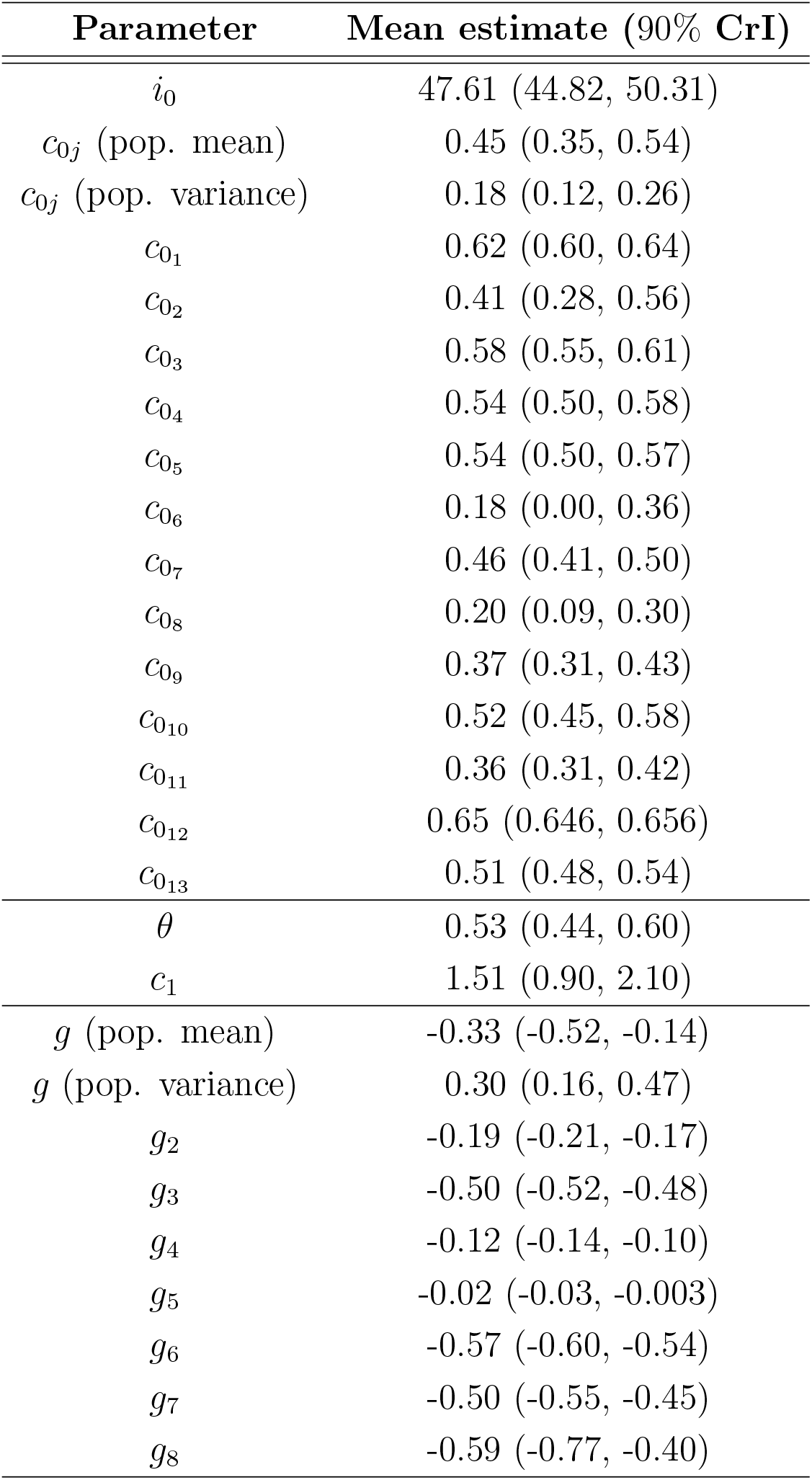
Estimated parameters for the distance matrix scenario. The estimated parameters obtained for the distance matrix scenario with 90% credible interval (CrI).

| Parameter | Mean estimate (90% CrI) |
| --- | --- |
| $i_0$ | 47.61 (44.82, 50.31) |
| $c_{0j}$ (pop. mean) | 0.45 (0.35, 0.54) |
| $c_{0j}$ (pop. variance) | 0.18 (0.12, 0.26) |
| $c_{01}$ | 0.62 (0.60, 0.64) |
| $c_{02}$ | 0.41 (0.28, 0.56) |
| $c_{03}$ | 0.58 (0.55, 0.61) |
| $c_{04}$ | 0.54 (0.50, 0.58) |
| $c_{05}$ | 0.54 (0.50, 0.57) |
| $c_{06}$ | 0.18 (0.00, 0.36) |
| $c_{07}$ | 0.46 (0.41, 0.50) |
| $c_{08}$ | 0.20 (0.09, 0.30) |
| $c_{09}$ | 0.37 (0.31, 0.43) |
| $c_{010}$ | 0.52 (0.45, 0.58) |
| $c_{011}$ | 0.36 (0.31, 0.42) |
| $c_{012}$ | 0.65 (0.646, 0.656) |
| $c_{013}$ | 0.51 (0.48, 0.54) |
| $\theta$ | 0.53 (0.44, 0.60) |
| $c_1$ | 1.51 (0.90, 2.10) |
| $g$ (pop. mean) | -0.33 (-0.52, -0.14) |
| $g$ (pop. variance) | 0.30 (0.16, 0.47) |
| $g_2$ | -0.19 (-0.21, -0.17) |
| $g_3$ | -0.50 (-0.52, -0.48) |
| $g_4$ | -0.12 (-0.14, -0.10) |
| $g_5$ | -0.02 (-0.03, -0.003) |
| $g_6$ | -0.57 (-0.60, -0.54) |
| $g_7$ | -0.50 (-0.55, -0.45) |
| $g_8$ | -0.59 (-0.77, -0.40) |

**Table S2:** Estimated parameters for the weekly mobility matrices scenario. The estimated parameters obtained for the scenario we used weekly mobility matrices with 90% credible interval (CrI).

| Parameter | Mean estimate (90% CrI) |
| --- | --- |
| $i_0$ | 50.19 (47.37, 53.04) |
| $c_{0j}$ (pop. mean) | 0.21 (0.05, 0.36) |
| $c_{0j}$ (pop. variance) | 0.35 (0.24, 0.47) |
| $c_{0_1}$ | 0.59 (0.56, 0.61) |
| $c_{0_2}$ | 0.25 (0.06, 0.45) |
| $c_{0_3}$ | 0.50 (0.47, 0.54) |
| $c_{0_4}$ | 0.38 (0.33, 0.43) |
| $c_{0_5}$ | 0.19 (0.08, 0.30) |
| $c_{0_6}$ | -0.30 (-0.54, -0.04) |
| $c_{0_7}$ | 0.01 (-0.10, 0.11) |
| $c_{0_8}$ | -0.19 (-0.36, -0.03) |
| $c_{0_9}$ | -0.02 (-0.15, 0.12) |
| $c_{0_{10}}$ | 0.34 (0.21, 0.45) |
| $c_{0_{11}}$ | 0.02 (-0.09, 0.15) |
| $c_{0_{12}}$ | 0.601 (0.597, 0.61) |
| $c_{0_{13}}$ | 0.37 (0.31, 0.43) |
| $\theta$ | 0.90 (0.72, 0.98) |
| $c_1$ | 2.11 (1.52, 2.69) |
| $g$ (pop. mean) | -0.28 (-0.45, -0.10) |
| $g$ (pop. variance) | 0.32 (0.19, 0.50) |
| $g_2$ | -0.16 (-0.17, -0.14) |
| $g_3$ | -0.48 (-0.50, -0.45) |
| $g_4$ | -0.08 (-0.10, -0.06) |
| $g_5$ | 0.02 (0.003, 0.04) |
| $g_6$ | -0.53 (-0.56, -0.50) |
| $g_7$ | -0.47 (-0.51, -0.43) |
| $g_8$ | -0.58 (-0.74, -0.40) |

### A.5 Regions and their population sizes

**Table S3:** List of regions and their population sizes. The thirteen local health areas (LHAs) in Fraser Health, British Columbia, Canada, and their population sizes. Source [6].

|  | Local Health Authority (LHA) | Population |
| --- | --- | --- |
| 1 | Abbotsford | 161,912 |
| 2 | Agassiz/Harrison | 10,770 |
| 3 | Burnaby | 257,926 |
| 4 | Chilliwack | 105,862 |
| 5 | Delta | 112,259 |
| 6 | Hope | 8,931, |
| 7 | Langley | 161,725 |
| 8 | Mission | 47,652 |
| 9 | Maple Ridge/Pitt Meadows | 111,502 |
| 10 | New Westminster | 82,590 |
| 11 | South Surrey/White Rock | 107,347 |
| 12 | Surrey | 512,436 |
| 13 | Tri-Cities | 254,583 |

### A.6 Results for the scenario with a fixed mobility matrix for the entire study period

In this section, we consider a scenario where mobility is described by fixed matrix computed from the Telus mobility data for the entire study period.

**Figure S9:**
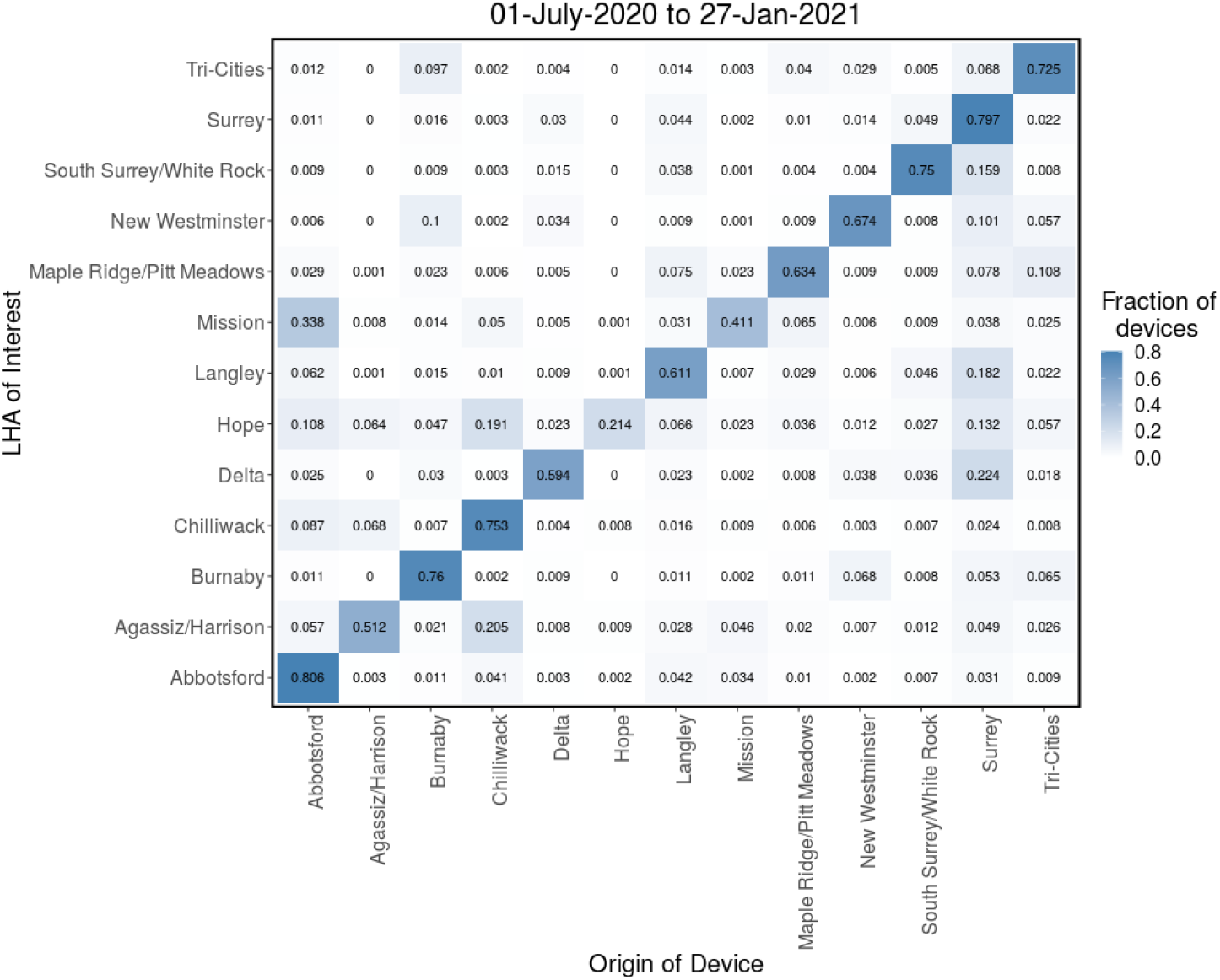
Fixed mobility matrix for the entire study period. Probability matrix (π) computed from the Telus mobility data for the entire study period from July 1, 2020 to Jan. 27, 2021. π_ji_ is the probability that an individual who migrated from one of the 13 LHAs to region j, originated from region i.

**Figure S10:**
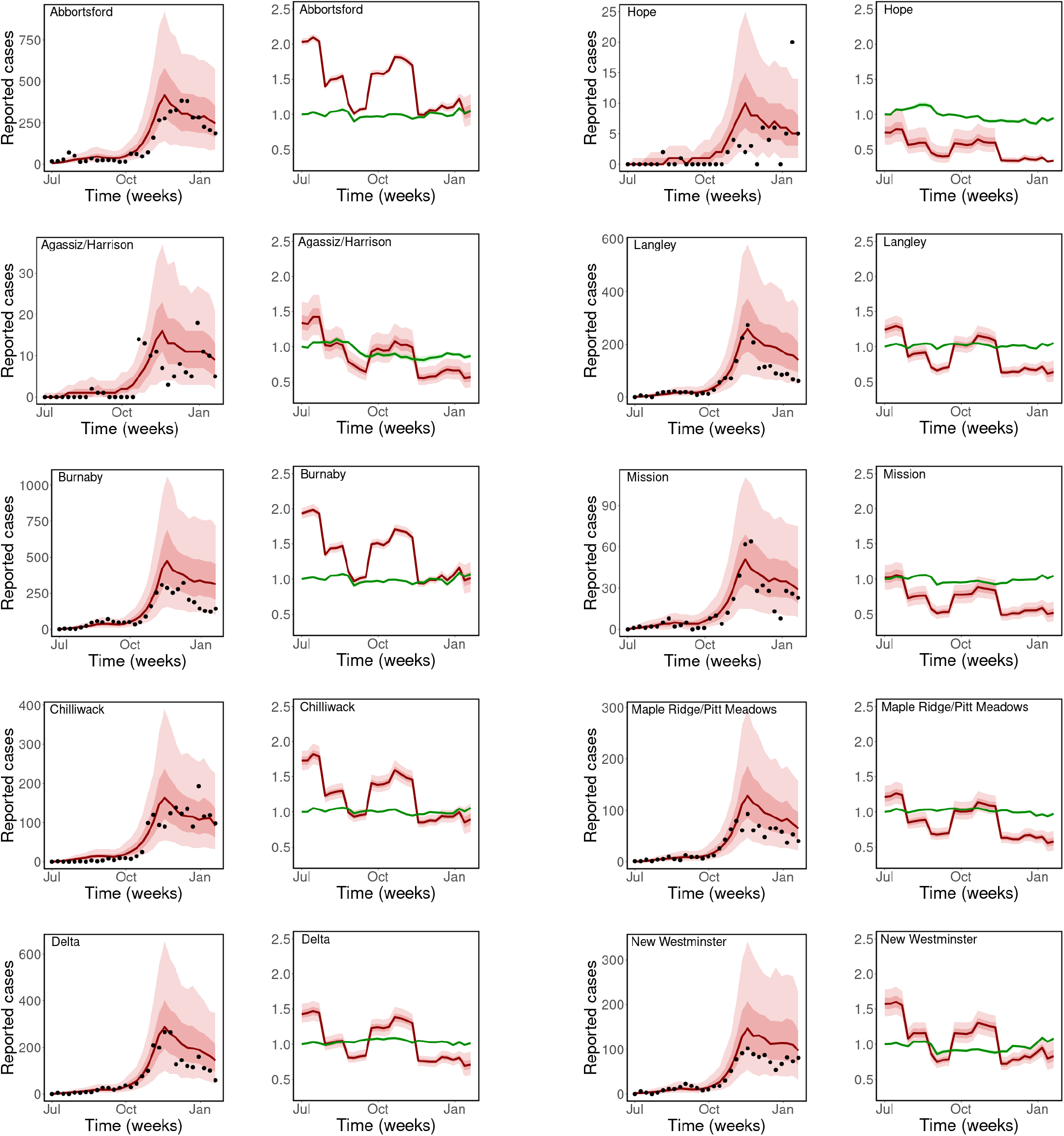
Observed and estimated COVID-19 cases. Weekly reported cases and model prediction (columns 1 and 3), computed using the fixed mobility matrix in Figure S9. Disease transmission rate, β_j_(t) and the contribution of mobility to disease transmission, 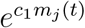 (columns 2 and 4). Black dots are the weekly reported cases of COVID-19, the solid lines are the mean estimates of cases/parameters, the darker bands are the 50% CrI, while the lighter bands are the 90% CrI.

**Figure S11:**
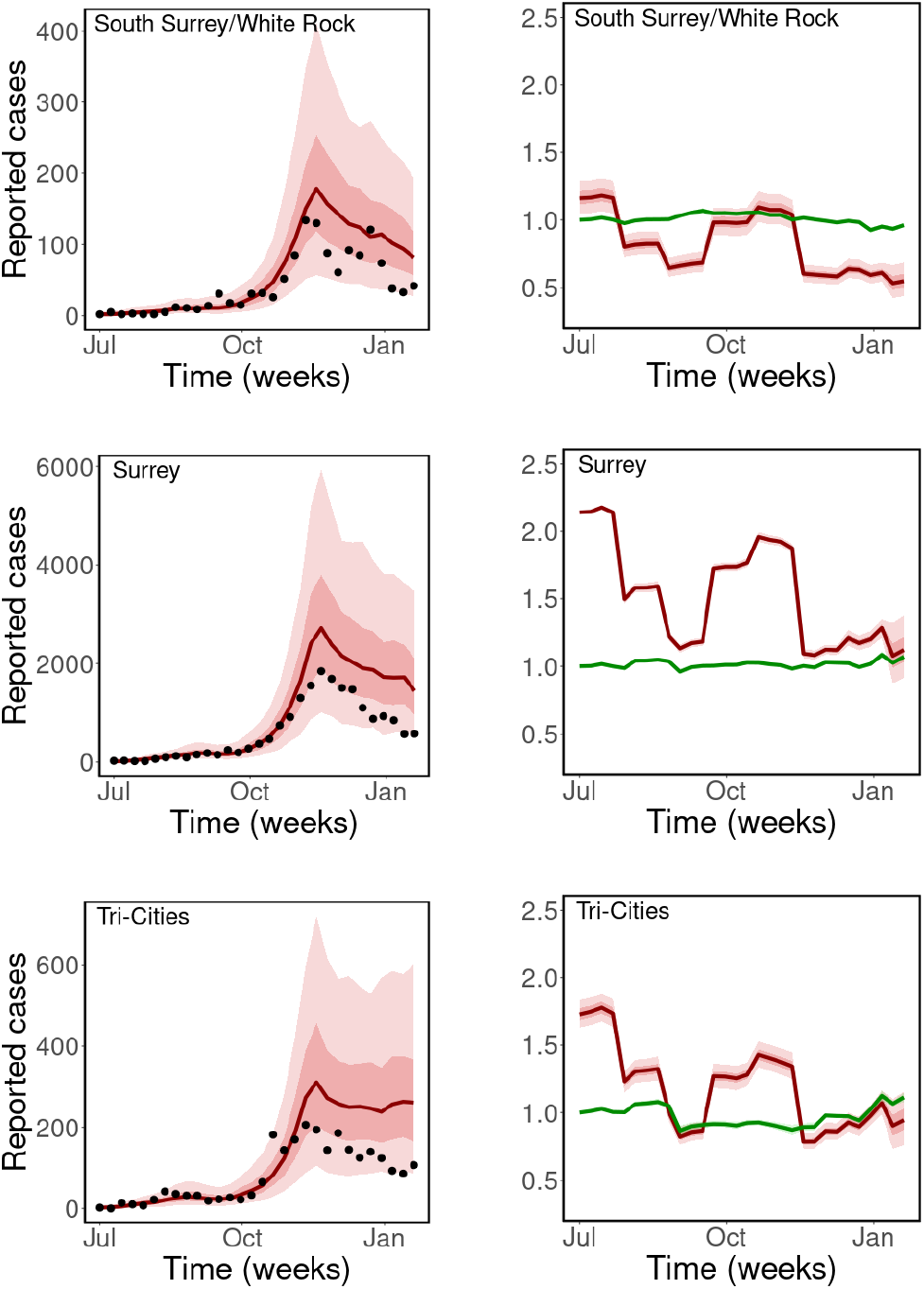
Observed and estimated COVID-19 cases. Weekly reported cases and model prediction (columns 1 and 3), computed using the fixed mobility matrix in Figure S9. Disease transmission rate, β_j_(t) and the contribution of mobility to disease transmission, 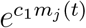 (columns 2 and 4). Black dots are the weekly reported cases of COVID-19, the solid lines are the mean estimates of cases/parameters, the darker bands are the 50% CrI, while the lighter bands are the 90% CrI.

**Table S4:** Estimated parameters for the fixed mobility matrix scenario. Estimated parameters with 90% credible interval (CrI).

| Parameter | Mean estimate (90% CrI) |
| --- | --- |
| $i_0$ | 35.75 (33.79, 37.78) |
| $c_{0j}$ (pop. mean) | 0.31 (0.14, 0.48) |
| $c_{0j}$ (pop. variance) | 0.38 (0.26, 0.52) |
| $c_{01}$ | 0.71 (0.69, 0.73) |
| $c_{02}$ | 0.30 (0.10, 0.50) |
| $c_{03}$ | 0.66 (0.62, 0.70) |
| $c_{04}$ | 0.54 (0.47, 0.63) |
| $c_{05}$ | 0.36 (0.26, 0.45) |
| $c_{06}$ | -0.29 (-0.56, 0.01) |
| $c_{07}$ | 0.21 (0.11, 0.32) |
| $c_{08}$ | 0.02 (-0.16, 0.18) |
| $c_{09}$ | 0.19 (0.07, 0.32) |
| $c_{010}$ | 0.45 (0.34, 0.58) |
| $c_{011}$ | 0.15 (0.05, 0.25) |
| $c_{012}$ | 0.76 (0.756, 0.77) |
| $c_{013}$ | 0.55 (0.49, 0.61) |
| $\theta$ | 0.60 (0.52, 0.70) |
| $c_1$ | 1.55 (1.10, 1.99) |
| $g$ (pop. mean) | -0.41 (-0.60, -0.21) |
| $g$ (pop. variance) | 0.31 (0.17, 0.48) |
| $g_2$ | -0.34 (-0.36, -0.32) |
| $g_3$ | -0.60 (-0.62, -0.57) |
| $g_4$ | -0.22 (-0.24, -0.20) |
| $g_5$ | -0.11 (-0.13, 0.10) |
| $g_6$ | -0.67 (-0.71, -0.64) |
| $g_7$ | -0.60 (-0.64, -0.55) |
| $g_8$ | -0.71 (-0.92, -0.51) |

